# Genome-wide association meta-regression identifies stem cell lineage orchestration as a key driver of acne risk

**DOI:** 10.1101/2025.06.27.25330406

**Authors:** Jessye Maxwell, Brittany L. Mitchell, Xinyi Du-Harpur, Luba M. Pardo, Willemijn C. A. M. Witkam, Nick Dand, Meike Bartels, Michael J Betti, Dorret I. Boomsma, Xianjun Dong, Zachary Gerring, Sarah Finer, Genes & Health Research Team, Fiona A Hagenbeek, Jouke Jan Hottenga, George Hripcsak, Laura Huilaja, Kristian Hveem, Benjamin M. Jacobs, Mart Kals, James Kaufman-Cook, Johannes Kettunen, Atlas Khan, Külli Kingo, Krzysztof Kiryluk, Mari Løset, Gerton Lunter, Michelle K Lupton, Josine L. Min, Nicholas G Martin, Sarah E Medland, Andres Metspalu, Dorien Neijzen, Tamar E. C. Nijsten, Tiit Nikopensius, Catherine M Olsen, Lynn Petukhova, Anu Reigo, Miguel E Rentería, Rossella Rispoli, Jake Saklatvala, Eeva Sliz, Kaisa Tasanen-Määttä, Maris Teder-Laving, Laurent Thomas, Richard C Trembath, Mariliis Vaht, David A van Heel, Chunhua Weng, David C Whiteman, Jonathan N. Barker, Catherine Smith, Michael A. Simpson

**Affiliations:** Department of Medical and Molecular Genetics, School of Basic and Medical Biosciences, King’s College London, London, United Kingdom; Brain and Mental Health program, QIMR Berghofer Medical Research Institute, Brisbane, Australia; School of Biomedical Sciences, Faculty of Health, Queensland University of Technology (QUT), Brisbane, Australia; School of Biomedical Sciences, Queensland University of Technology, Brisbane, Australia; St John’s Institute of Dermatology, School of Basic and Medical Biosciences, Faculty of Life Sciences and Medicine, King’s College London, London, United Kingdom; Generation R Study Group, Erasmus MC, University Medical Center Rotterdam, Rotterdam, the Netherlands; Department of Dermatology, Erasmus MC, University Medical Center Rotterdam, Rotterdam, the Netherlands; Department of Biological Psychology, Vrije Universiteit Amsterdam, Amsterdam, the Netherlands; Amsterdam Public Health Research Institute, Amsterdam, the Netherlands; Department of Medicine, Division of Genetic Medicine, Vanderbilt University Medical Center, Nashville, TN, USA; Department of Complex Trait Genetics, Center for Neurogenomics and Cognitive Research, Vrije Universiteit Amsterdam, Amsterdam, the Netherlands; Amsterdam Reproduction & Development (AR&D) research institute, Amsterdam, the Netherlands; Genomics and Bioinformatics Hub, Brigham and Women’s Hospital, Boston, MA, USA; Department of Neurology, Brigham and Women’s Hospital, Boston, MA, USA; Adams Center of Parkinson’s Disease Research, Department of Neurology, Yale School of Medicine, Yale University, New Haven, CT, USA; Walter and Eliza Hall Institute of Medical Research, VIC, Australia; Wolfson Institute of Population Health, Queen Mary University of London, London, UK; Institute for Molecular Medicine Finland (FIMM), HiLIFE, University of Helsinki, Helsinki, Finland; Department of Biomedical Informatics, Vagelos College of Physicians and Surgeons, Columbia University, New York, NY, USA; Research Unit of Clinical Medicine, University of Oulu, Oulu, Finland; Medical Research Center, Oulu University Hospital, Oulu, Finland; HUNT Center for Molecular and Clinical Epidemiology, Department of Public Health and Nursing, NTNU - Norwegian University of Science and Technology, Trondheim, Norway; HUNT Research Centre, Department of Public Health and Nursing, Norwegian University of Science and Technology, Levanger, Norway; Levanger Hospital, Nord-Trøndelag Hospital Trust, Levanger, Norway; Estonian Genome Center, Institute of Genomics, University of Tartu, Tartu, Estonia; Systems Epidemiology, Research Unit of Population Health, Faculty of Medicine, University of Oulu, Oulu, Finland; Division of Nephrology, Department of Medicine, Vagelos College of Physicians and Surgeons, Columbia University, New York, NY, USA; Faculty of Medicine, Institute of Clinical Medicine, University of Tartu, Tartu, Estonia; Tartu University Hospital, Tartu, Estonia; Department of Dermatology, Clinic of Orthopaedy, Rheumatology and Dermatology, St. Olavs Hospital, Trondheim University Hospital, Trondheim, Norway; Department of Epidemiology, University of Groningen, University Medical Center Groningen, Groningen, the Netherlands; MRC Integrative Epidemiology Unit, University of Bristol, Bristol, UK; Population Health Sciences, Bristol Medical School, University of Bristol, Bristol, UK; Psychiatric Genetics, QIMR Berghofer Medical Research Institute, Brisbane, Australia; School of Psychology, University of Queensland, Brisbane, Australia; School of Psychology and Counselling, Queensland University of Technology (QUT), Brisbane, Australia; Population Health program, QIMR Berghofer Medical Research Institute, Brisbane, Australia; The Ronald O. Perelman Department of Dermatology, NYU Grossman School of Medicine, NYU Langone Health, New York, New York, USA; Department of Population Health, NYU Grossman School of Medicine, NYU Langone Health, New York, New York, USA; Department of Clinical and Molecular Medicine, NTNU - Norwegian University of Science and Technology, Trondheim, Norway; BioCore - Bioinformatics Core Facility, NTNU - Norwegian University of Science and Technology, Trondheim. Norway; Blizard Institute, Barts and the London School of Medicine and Dentistry, Queen Mary University of London, London, UK

## Abstract

Over 85% of the population experience acne at some point in their lives, with its severity spanning a quantitative spectrum, from mild, transient outbreaks to more persistent, severe forms of the condition. Moderate to severe disease poses a substantial global burden arising from both the physical and psychological impacts of this highly visible condition.

The analytical approach taken in this study aimed to address the impact of variation in the dichotomisation of acne case control status, driven by ascertainment and study design, on effect size estimates across independent genetic association studies of acne. Through a fixed intercept meta-regression framework, we combined evidence genome-wide for association with acne across studies in which case-control status had been ascertained in different settings, allowing for different severity threshold definitions. Across a combined sample of 73,997 cases and 1,103,940 controls of European, South Asian and African American ancestry we identify genetic variation at 165 genomic loci that influence acne risk. There is evidence for both shared and ancestry specific components to the genetic susceptibility to acne and for sex differences in the magnitude of effect of risk alleles at three loci.

We observe that common genetic variation explains 13.4% of acne heritability on the liability scale. Consistent with the hypothesis that genetic risk primarily operates at the level of individual pilosebaceous units, a polygenic score derived from this case-control study of acne susceptibility is associated with both self-reported and clinically assessed acne severity in adolescence, further strengthening the link between genetic risk and disease severity.

Prioritisation of causal genes at the identified acne risk loci, provides genetic validation of the targets of established and emerging acne therapies, including retinoid treatments. The identified acne risk loci are enriched for genes encoding downstream effectors of *RXRA* signalling, including *SOX9* and components of the *WNT* and p53 pathways. Illustrating that the control of stem cell lineage plasticity and cellular fate are important mechanisms through which genetic variation influences acne susceptibility within the pilosebaceous unit.

## Introduction

Acne is the most common human skin disease. It typically emerges during adolescence and affects nearly all teenagers to varying degrees.^1^ The characteristic lesions of the pilosebaceous unit, including comedones, papules, and pustules, primarily manifest during this age range, but for a substantial subset of individuals, the condition persists into adulthood.^1^

The severity of acne varies widely. Approximately 16% of the population experience moderate to severe acne^1^ and over 20% of these individuals develop long-term scarring. Beyond its physical manifestations, acne is associated with a significant mental health burden, highlighting the profound psychological impact of this highly visible condition.^2^ The societal consequences of acne are substantial, contributing to an estimated 5 million Disability-Adjusted Life Years (DALYs) globally.^3^ Acne is observed in all global populations, though prevalence estimates vary, with higher rates reported in economically developed and socially advantaged regions.^4^ Age standardised rates of disease have increased globally by 0.55% per year over the last 30 years, with faster increases in males that have narrowed the sex difference in prevalence, which is currently estimated to be 1.3 times higher in females.^4^

There is an unmet need for therapies that are both highly effective and well-tolerated for the treatment of moderate to severe acne. Current treatments, including topical medications, systemic antibiotics, and hormonal therapies, often fail to adequately resolve severe disease.^5^ Conversely, Isotretinoin, a retinoid derived from vitamin A, is highly effective, but its use is constrained by its adverse effect profile, including mucocutaneous symptoms, mood alterations, and severe teratogenic effects, which necessitate close monitoring and strict precautions to prevent pregnancy.^6^ These limitations underscore the pressing need for new therapeutic approaches that not only address the underlying causes of acne but also minimise associated risks.

Improved understanding of the genetic susceptibility to acne offers opportunity to identify the causal molecular and cellular mechanisms of the disease. An effective treatment strategy for acne must not only aim to reduce the time to healing of existing lesions, but also prevent naive pilosebaceous units entering an acne cycle. Even in areas of severely affected skin < 0.5% of pilosebaceous units are involved in visible lesions and the mean lifetime of each lesion is of the order of days for inflammatory lesions and weeks for non-inflammatory lesions. Given that only a small fraction of pilosebaceous units are affected at any one time, it is likely that acne risk factors operate at the level of individual pilosebaceous units, influencing their propensity to develop into acne lesions. Genetic variation plays a substantial role in acne risk, with heritability estimates reaching up to 80%.^7–11^ Recent genome-wide association studies (GWAS) have identified genetic variation at 49 genomic loci that contribute to acne risk.^12–16^ A polygenic risk score for acne has been shown to correlate with self-reported disease severity,^12^ consistent with a relationship between genetic liability to acne and the seveerity of disease manifestation.

To deepen our understanding of the genetic architecture of acne, we have undertaken and interrogated large-scale genome-wide association studies for combined and sex-specific effects across 18 individual studies, including participants from diverse ancestry groups. We integrated evidence of association across studies while accounting for differences in the dichotomisation of acne, which may be considered a quantitative trait, in different ascertainment settings. We identify genetic variation influencing acne risk at 117 novel risk loci. Prioritisation of putative causal genes and cell types provide genetic support for both established and emerging acne therapies and implicates variation in the molecular control of lineage plasticity and cellular fates in the pilosebaceous unit as an important contributor to acne susceptibility.

## Results

### Genome-wide association meta-analysis in populations of European ancestry

We performed genome-wide association testing of common genetic variants on autosomes and the X chromosome with acne, analysing data from 16 independent case-control studies comprising 69,333 cases and 1,051,871 controls of European ancestry (cohort details in ***Supplementary Note & Table S1***). Ascertainment methods varied across the 16 cohorts, ranging from clinical diagnoses of acne vulgaris by dermatologists to self-reported measures. Effect size estimates of lead variants at previously reported acne risk loci were compared across all 16 studies, which revealed evidence of systematic heterogeneity in effect size estimates between cohorts. We noted that cohorts in which acne cases had been specifically recruited in clinical settings, and thus likely representing individuals with more severe acne, contributed consistently larger effect size estimates than those relying on electronic health records or self-reported data (***Supplementary Figure 1***). We hypothesised that these systematic differences could be explained in part by different approaches to ascertainment and study design equating to different effective thresholds in severity and underlying liability distributions of cases and controls.

Capitalising on quantitative genetic theory that links the observed scale heritability and underlying liability scale for threshold selected binary traits,^17^ we utilised a simple heuristic approach to estimate the difference in the mean liability between case and control participants in each study that are implied by the ascertainment procedures of each constituent study (***Figure 1a*** and ***Methods***). The resulting parameter (difference in mean liability Δ_L_) explained 88% of the variance in effect size differences observed at established acne risk loci across the 16 cohorts (***Figure 1b***). To account for this ascertainment-induced bias whilst combining evidence of association across the 16 independent studies, we performed a fixed intercept meta-regression (FIMR) weighted by the study-level parameter Δ_L_ to adjust for ascertainment and an intercept fixed at zero. The estimated effect and corresponding standard error of each variant were extracted from the FIMR model to correspond to an acne prevalence of 16%, equivalent to a case control study of moderate to severe acne.

**Figure 1.**
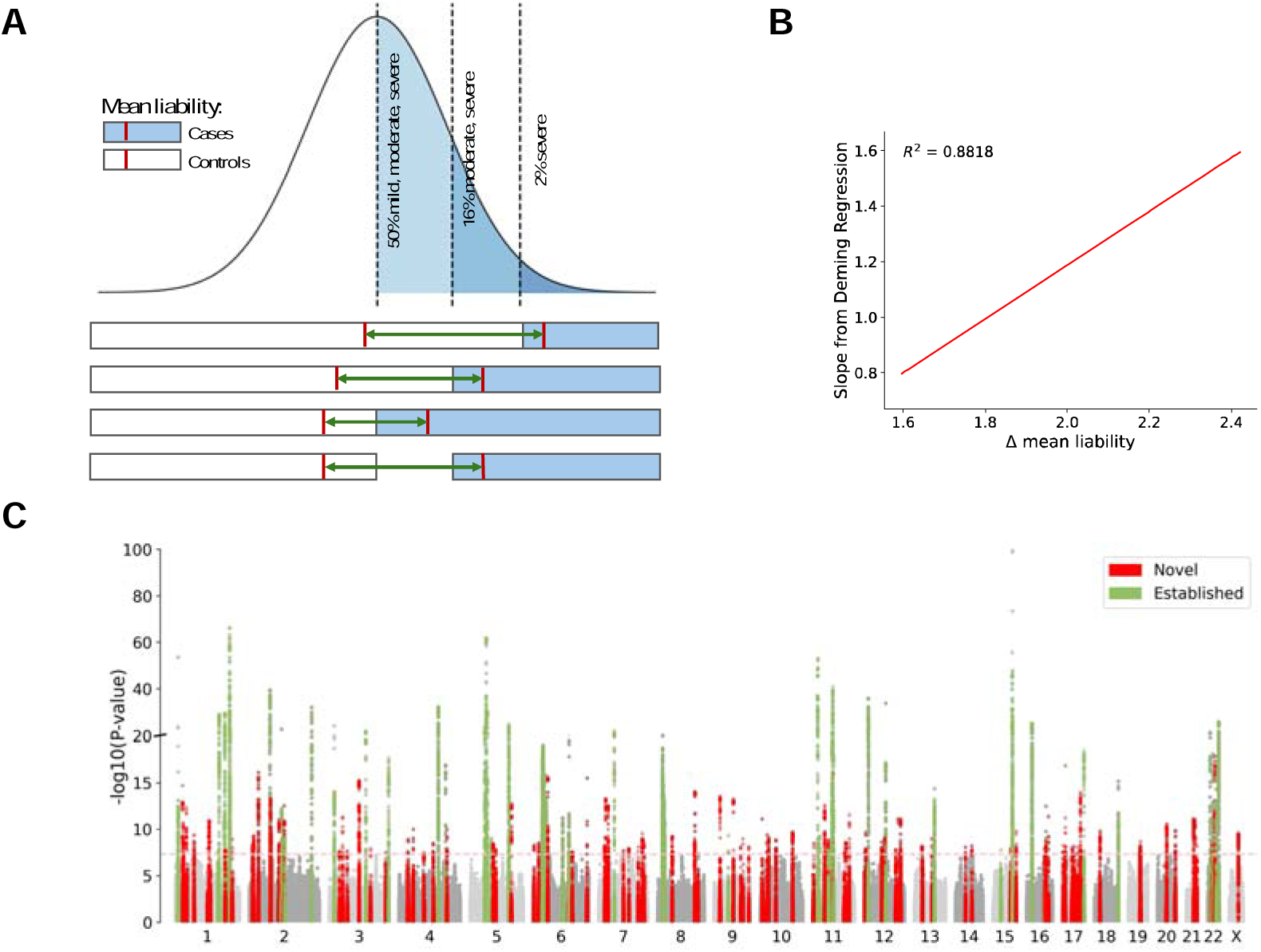
A) Approach to estimate ascertainment effects from the capture of different severity classes. Prevalence estimates of acne severity classes from epidemiological studies overlaying a normal distribution representing the acne liability distribution, with 50% of the population having a history of mild, moderate or severe acne, 14% moderate or severe, and 2% severe. Below the distribution, illustrates how the study-level parameter is determined for different ascertainment approaches. The areas representing cases are shaded in blue, while controls are shaded in white. The red bar indicates the mean liability for cases and controls. The difference in mean liability between cases and controls, which defines the ascertainment study-level parameter, is represented by the double headed green arrows. **B) Systematic differences in effect sizes distributions for each of the 16 studies against the study’s respective ascertainment study-level parameter.** The size of each point proportional to the sample size of the respective study. Systematic differences in effect sizes distributions for each study is measured by the slope of a Deming regression of the study’s effect size estimates on the fixed effect meta-analysis effect estimates of the lead variants at established acne loci (Methods). The R-squared (R^2^) value represents the proportion of variance in the systematic differences in effect sizes explained by the study-level parameter. **C) Evidence of association of genetic variants across the genome with acne.** Genetic variants ordered by chromosome and position along the x-axis with the evidence of association, −log10(P-value) from the association analysis using a two-sided Z-test, on the y-axis. The P-values are not adjusted for multiple comparisons, but the dashed pink line marks the genome-wide significance threshold (P = 5 × 10^−8^). Green points represent variants at established acne susceptibility loci, while points represent variants at novel risk loci.

Inflation of test statistics across the genome was observed in the FIMR at a similar level to those arising from a fixed effect meta-analysis (FEMA) model (*λ_GC_* = 1.393 and 1.386, respectively). LD score regression intercepts (*I*_LDSC_ = 1.0956 (s.e. = 0.011) and 1.0944 (s.e. = 0.0114), respectively) also indicate inflation of test statistics, though the attenuation ratios (R_PS_ = 0.1386 (s.e. = 0.0167) and 0.1436 (s.e. = 0.0166), respectively) suggest that this inflation is principally driven by the polygenicity of acne combined with the large sample size of the study, rather than systematic bias or population stratification.^18^

Across the genome common genetic variants assessed for association using the FIMR model explained 13.4% (s.e. = 0.00894) of the variance in liability of acne, with 14,612 variants surpassing genome-wide significant evidence of association compared to 13,996 observed with FEMA model. These association signals represent 296 and 290 LD independent (r2 < 0.1) lead variants that are spread across 155 and 157 LD independent blocks^19^ respectively. The majority of these loci were common to both methods, with only five and seven unique to either the FIMR approach or the FEMA respectively. Across the 150 susceptibility loci, harbouring variants with genome-wide significant effects under both models, there is strong evidence that the FIMR model provides an improved fit to the data compared to the FEMA model. The distribution of FIMR model deviance measures across the lead variants of these 150 identified risk loci are lower than those arising from the FEMA model (p=5.47×10^−7^; detailed comparisons of the results from the FIMR and FEMA models are provided in ***Supplementary Table S2)***. Genome-wide significant association signals were observed at 48 of the 49 previously reported acne susceptibility loci (***Figure 1c***; ***Supplementary Table S2***).^12–15^ The only previously reported risk locus at which no genome-wide significant evidence of association was observed at 1q24.2, which was previously identified as an acne risk locus in a Han Chinese population (previously reported lead variant rs7531806; OR=0.99; 95%CI 0.98-1.01; P=0.46).^13^

A key advantage of modelling the differences in ascertainment with the FIMR approach should be better calibration of estimated allelic effect sizes on acne risk. To investigate the calibration of effect sizes across the genome, we generated a series of polygenic risk scores (PRS) from the GWAS summary statistics from both the FIMR and FEMA models. We evaluated the performance of the risk scores in an independent cohort of 2,058 individuals for whom the history of acne had been evaluated by questionnaire. The PRS generated by both approaches all had strong evidence of association with acne in this independent cohort, with the point estimates of the effect sizes of PRS generated from the FIMR consistently larger than those generated from the FEMA (***Supplementary Table S3***). The strongest evidence of association was observed with the PRS derived from the FIMR using the SBayesR method, which leverages genome-wide effects across all SNPs (beta=0.078, SE=0.010, p=1.75×10^−15^), corresponding to an estimated 3.5% of acne liability explained compared to an estimated 2.7% with a PRS derived from the previous largest GWAS meta-analysis of acne.^12^

Consistent with previous observations, the mean of the PRS distribution significantly increases with self-reported disease severity (***Figure 2***), with higher PRS corresponding to higher likelihood of reporting more severe disease (none, mild, moderate, severe). An ordinal regression between acne severity groups showed that the risk of a single category increase in acne severity increased by 1.63-fold for every one standard deviation increase in PRS (OR=1.63, 95% CI 1.45-1.73; P=6.83×10^−16^). To further evaluate this relationship between genetic liability and disease severity and overcome the potential limitations of self-reported evaluation of disease severity we investigated the distribution of PRS in a cross-sectional cohort of 2,482 teenagers. Within this cohort acne severity was ascertained and categorised (clear/almost clear, mild, moderate/severe) by trained dermatologists from three-dimensional facial photographs taken at age ∼13.^21^ Consistent with the findings with self-reported acne, a single standard deviation increase in polygenic risk corresponds to a 1.53-fold increase in the odds of being one severity category higher (OR = 1.53, 95% CI 1.41-1.65; P = 2.68×10^−26^).

**Figure 2:**
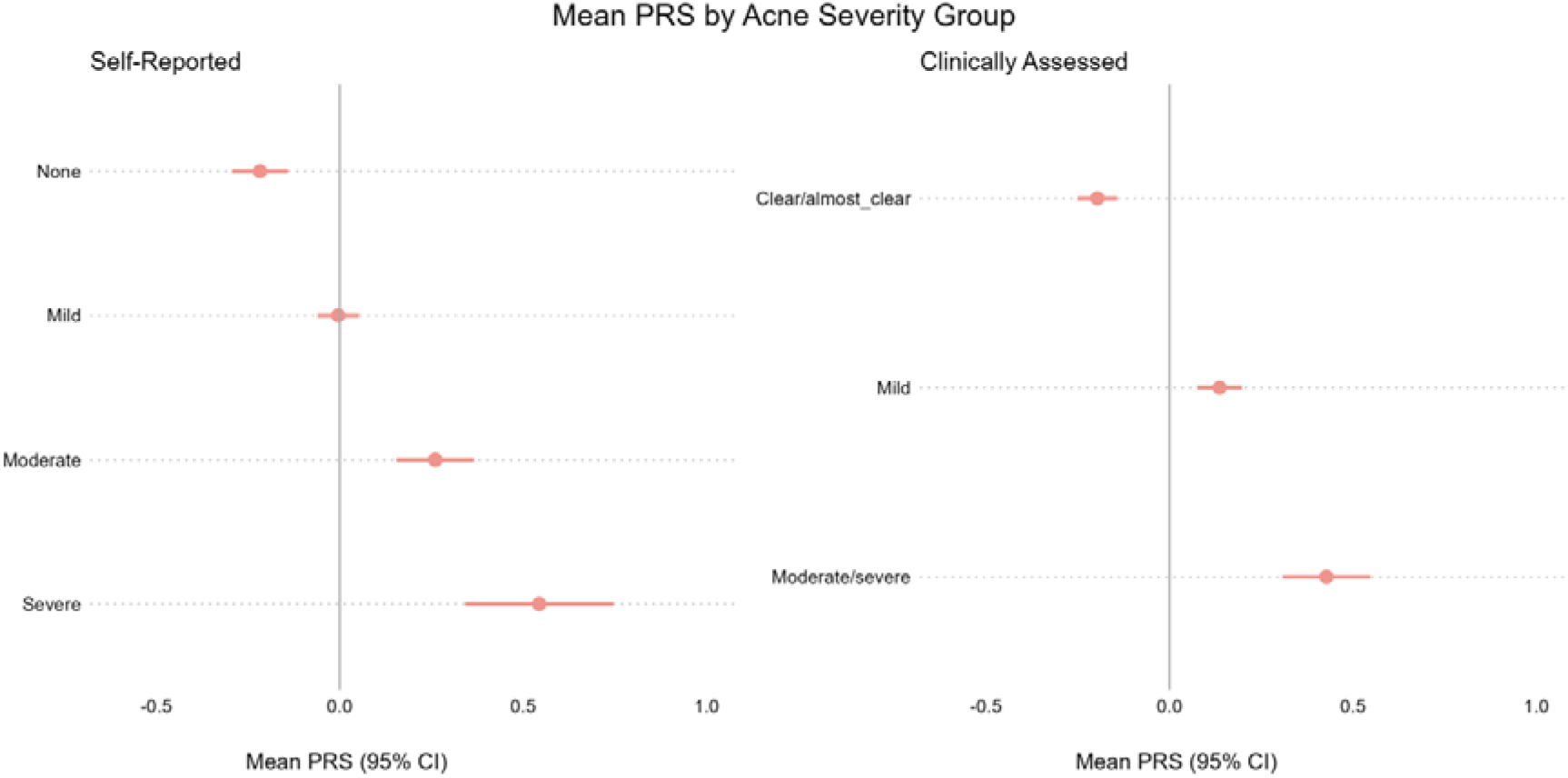
Mean of the polygenic risk score distributions by acne severity classifications in two independent cohorts. The left-hand panel shows the PRS distributions in the PISA cohort, in which acne severity was assessed using a self-reported retrospective questionnaire, the right-hand panel the Generation R cohort, where acne severity was assessed by trained dermatologists. The x-axis denotes the mean PRS calculated using SBayesR from the FIMR results and the y-axis represents acne severity classification in each cohort. The bars demark the 95% confidence intervals for the mean.

### Evaluation of the consistency of genetic susceptibility to acne across global populations

The genetic basis of acne has only previously been investigated in populations with recent European and Han Chinese ancestry.^12–15^ We evaluated genome-wide evidence of genetic association with acne in two cohorts from previously unstudied populations; a UK population cohort comprising individuals of South Asian (Pakistani and Bangladeshi) heritage with 4,094 acne cases and 37,935 controls and an African American cohort comprising 570 cases and 14,134 controls.

Of the 162 acne susceptibility loci identified in European populations, only one is observed with genome-wide significant evidence of association with acne in either cohort; 5q11.2 (rs38056; OR = 1.16; 95% CI 1.10–1.23; *P* = 1.5×10^−8^) in the South Asian study. Whilst the asymmetry of sample sizes across these populations will impact power to identify associations of the magnitude observed in the European ancestry meta-analysis, across the lead variants at each of the 162 acne susceptibility loci only 22 had evidence of a divergent effect size in the South Asian study compared to Europeans (***Supplementary Table S4)*** and seven in the African American study compared to Europeans (***Supplementary Table S5***). Notably, the risk allele frequencies of 18 of these 29 variants with divergent effects are >10% between the respective populations.

In the African American study, a genome-wide significant association was observed at 3q25.33 (rs2243130; OR = 2.27; 95% CI 1.90-2.73; P = 5×10^−9^). This variant, which is located in the third intron of *IL12A*, is not associated with acne in the European FIMR (rs2243130, P = 0.65), nor is there any genome-wide significant evidence of association of any variant within 500kb of rs2243130. However, there is nominal evidence of association and consistent direction of effect of the lead variant in the South Asian study (rs2243130; OR = 1.32, 95% CI 1.06-1.65; P = 0.02).

### Investigation of sex-specific effects on acne susceptibility

Acne exhibits sexual dimorphism with variation in affected areas, duration of disease and fluctuation in symptoms that may relate to the menstrual cycle in females. To identify genetic variation that influences susceptibility to acne differently between males and females, we undertook sex stratified GWAS FIMR across the 16 case-control studies of European ancestry participants. The female analyses comprised a total of 47,259 cases, 582,604 controls and the male analysis 22,066 cases, 469,137 controls. We observed genome-wide significant evidence of association of genetic variation with acne at 78 loci in females and 46 loci in males. These included the 7q21.2 locus in males and 5q13.3 in females, which were not observed at genome-wide significance in the combined analysis. The lead variant at 7q21.2 in males (rs2974116; OR=1.07; 95% CI 1.05–1.10; P=1.46×10^−8^) exhibits nominally significant evidence of association with acne in females (rs2974116, OR= 1.02; 95% CI 1.00–1.04; P=0.014), but with a significant difference in effect sizes between sexes (rs2974116, P*_int_*=0.0015). At 5q13.3 the lead variant in females (rs11744445, OR = 1.04; 95% CI 1.03–1.06; P 2.47×10^−8^) was non-significant in males (rs11744445, OR = 1.01; 95% CI 0.99–1.03; P = 0.46) with a significant difference in these effect estimates (rs11744445; P*_int_*= 0.009, ***Figure 3, Supplementary Table S6***).

**Figure 3.**
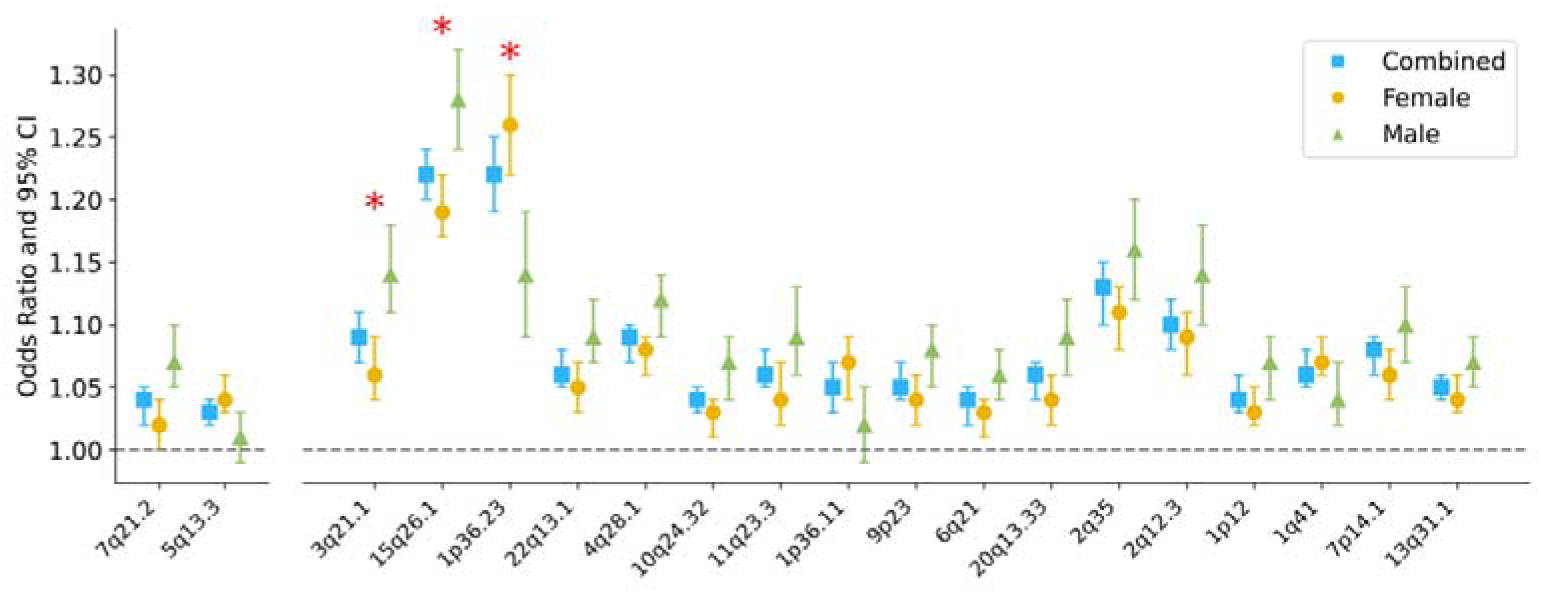
Sex specific genetics effects on acne risk. Odd ratios of the allelic effects on acne risk in males, females and combined analysis. The two variants on the left are those identified with a genome-wide significant effect on acne risk in the sex stratified analyses, and the 17 on the right are the lead variants from the 162 acne risk loci identified in the combined analysis for which there is a nominally significant difference in magnitude between male and female effect sizes (P<0.05, two-sided Z-difference test). The x-axis lists the chromosome band where the lead variant is located, and the y-axis represents the odds ratios. The bars with demark the 95% confidence intervals. Variants whose evidence of a difference in effect between males and females surpasses a Bonferroni-adjusted significance threshold for 162 tests (P<3.08×10^−4^) are marked with a red asterisk.

We examined whether any of the allelic associations at the 162 acne susceptibility loci identified via FIMR or FEMA in the combined European ancestry analysis displayed evidence of effect size differences between males and females. The lead variants at three of the 162 risk loci exhibited sex bias after adjusting for multiple testing (P<3.08×10^−4^) (***Supplementary Table S7***). In all three cases, there is evidence of an effect on acne risk in both males and female, but with a difference in the magnitude of effect; two (at 3q21.1 and 15q26.1) with a larger effect estimate in males and the third (at 1p36.23) where the larger effect was observed in females (***Figure 3, Supplementary Table S7***). The lead variants at a further 14 acne susceptibility loci demonstrate nominally significant (*P* < 0.05) evidence of a differential effect across sexes. This includes 2q35, at which the p. Phe228Ile (rs121908120) missense variant in *WNT10A* has been previously reported to have a larger effect in males. rs121908120 is not the lead variant at this locus in the current study, though it does demonstrate evidence consistent with a larger effect on acne risk in males (rs121908120, P*_int_* = 0.001).

### Identification of putative causal genes at acne susceptibility loci

Across the combined total of 165 genomic loci at which genetic variation with acne has been robustly associated with acne in this study, we sought to identify putative causal genes through which the allelic effects are likely impacting the biological mechanisms of disease susceptibility. We considered evidence from a range of sources linking acne associated variants to specific genes, and biological relationships between genes at distinct susceptibility loci (methods).

Evidence that acne associated genetic variation influences the expression of specific genes in skin was assessed by both colocalisation (***Supplementary Table 8)*** and imputation of expression levels using a transcriptome-wide association study (TWAS) framework (***Methods, Supplementary Table 9***). Skin was selected as the most appropriate available tissue for this analysis, supported by an investigation of the enrichment of acne association signals in proximity to genes expressed across 205 tissues and cell types using partitioned LD-score regression (***Supplementary Figure 2*** & ***Supplementary Table 10***, sun exposed skin *P* = 5×10^−7^; not sun exposed skin *P* = 8×10^−7^). The colocalisation and TWAS analysis identified 23 and 61 genes where there is evidence that expression in skin is influenced by acne susceptibility variants, with expression of 21 genes implicated by both methods.

We identified a further 161 putative causal genes in which the lead variant (or a variant in high LD, r^2^ > 0.8) of the acne association signal disrupts the protein coding sequence, an essential splice site or an established enhancer whose activity is predicted to influence expression of a specific gene (32 missense variants, 2 nonsense variants, 1 donor splice site variant and 136 variants within enhancer sequences, ***Supplementary Tables 11 and 12***).

Additionally, we prioritised genes with evidence of shared biological roles using established bioinformatic tools (DEPICT, ***Supplementary Table 13*** and Polygenic Priority Score (PoPs) ***Supplementary Table 14***). Rationalisation of this list of genes with links to acne associated genetic variation or shared biological roles across susceptibility loci, and selection of the nearest gene to the lead variant at loci where no genes with shared biological relationships were observed, resulted in a total of 243 genes across 165 loci that were prioritised for downstream investigations (***Figure 4*** & ***Supplementary Table 15)***. This list included genes previously implicated in acne biology at established risk loci, including *WNT10A, SEMA4B, LAMC2, IL36RN and TGFB2*.

**Figure 4:**
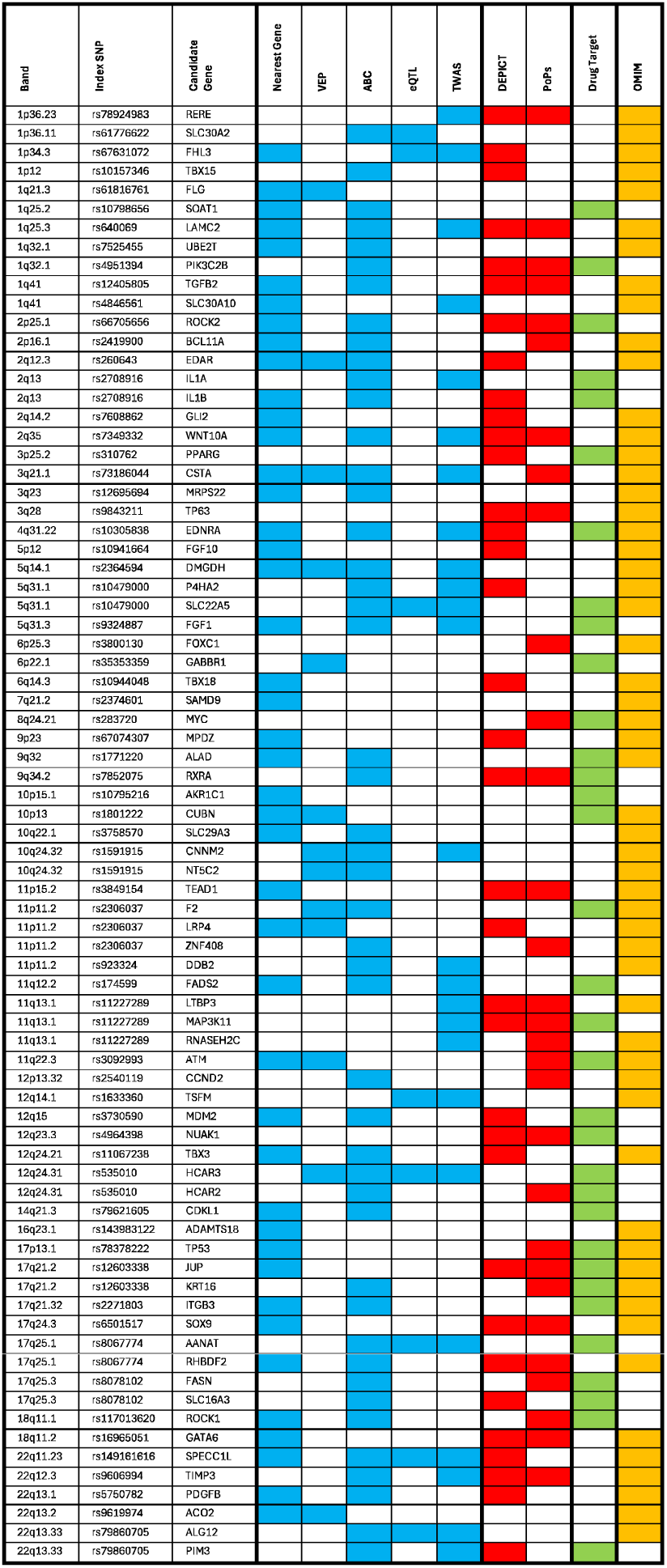
Prioritisation of candidate causal genes that are disrupted by rare alleles that cause Mendelian disorders or encode therapeutic targets. Genes prioritised by local evidence are shown in blue, encompassing nearest gene to the lead variant of the association signal, coding, splice site or enhancer variants in high LD (r2 > 0.8) with the lead variant, eQTL colocalization and significant TWAS associations in sun exposed or not sun exposed skin. Red boxes indicate gene prioritised by methods that examine shared biological functions across loci with DEPICT and Polygenic Priority Score (PoPs). The 58 genes represented in the table are all located at acne susceptibility loci identified in this study for the first time and encode the molecular target of an approved drug therapy (green) and are disrupted by rare alleles in Mendelian disorders (yellow).

### Identification of cell types and biological pathways in acne susceptibility

To identify cell types through which associated genetic variation may influence an individual’s susceptibility to acne, we clustered the set of 243 putative causal genes on the basis of expression levels in facial skin cell-types (***Methods***, ***Supplementary Figure 3***). This revealed populations of facial skin keratinocytes, pericytes and fibroblasts in which subsets of the putative causal genes are highly expressed. To investigate the biological processes that these genes may influence in these cell types, we clustered the genes on the basis of established direct and indirect gene-gene relationships (***Figure 5, Methods***) and noted the enrichment of an interacting group of genes that are highly expressed in keratinocytes in facial skin and contribute to keratinocyte differentiation (p=2.81×10^−10^) and formation of the cornified envelope (p=6.6×10^−17^). Other interacting gene modules are enriched for growth factor activity, Wnt signalling, cell adhesion and p53 signalling pathways (***Figure 5***, ***Supplementary Table 16***).

**Figure 5.**
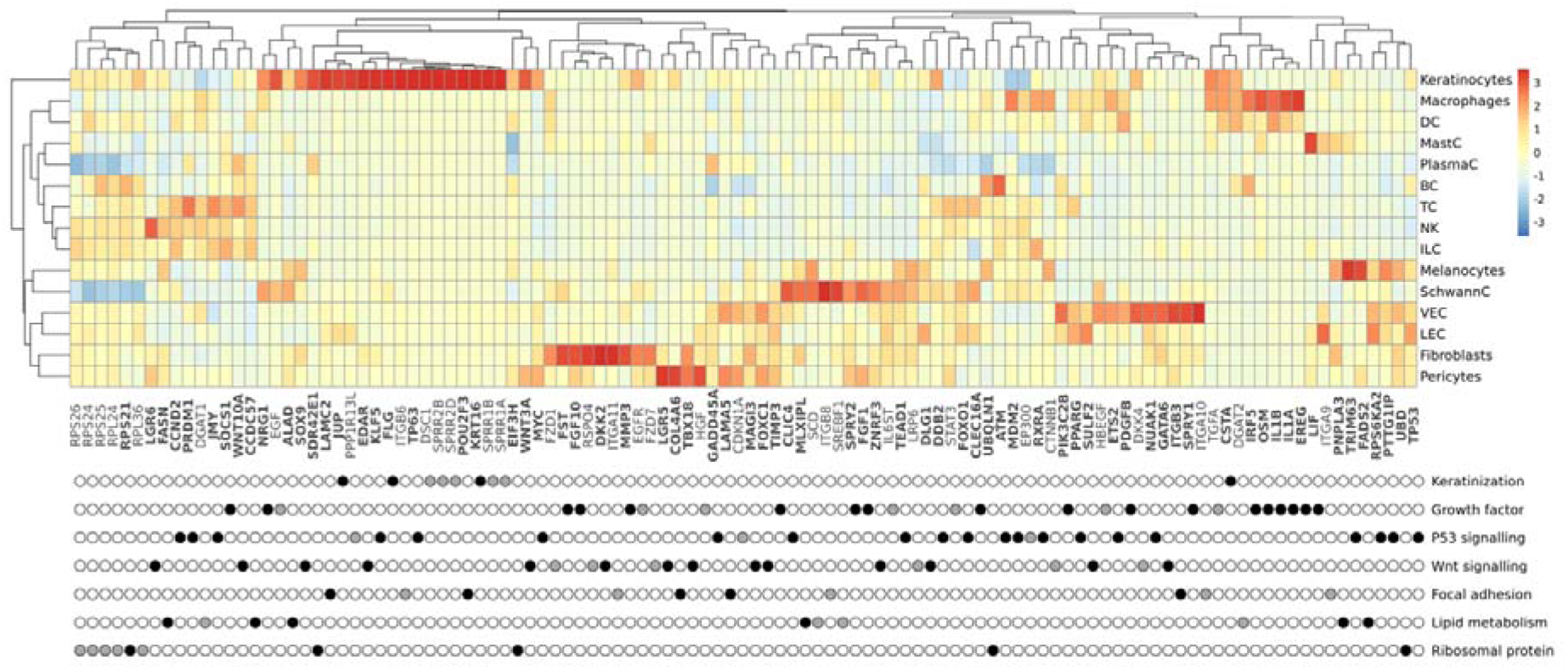
Clustering of expression levels of selected genes across different cell types in facial skin and annotation of gene clusters. The heatmap indicates scaled transcript levels within cell types: Keratinocytes, Macrophages, Dendritic Cells (DC), Mast Cells (MastC), Plasma Cells (PlasmaC), B Cells (BC), Innate Lymphoid Cells (ILC), T Cells (TC), Natural Killer cells (NK), Vascular Endothelial Cells (VEC), Lymphatic Endothelial Cells (LEC), Schwann Cells (SchwannC), Pericytes, Fibroblasts, and Melanocytes. Each row of the bottom plot indicates genes that have been clustered using evidence of direct and indirect (where a single intermediary gene serves as a linking bridge) gene-gene relationships from the STRING database. Clusters have a minimum size of four genes located across four acne loci. Black points indicate the prioritised putative causal genes identified at acne susceptibility loci, while grey points the intermediary genes in each cluster. Descriptions of the gene clusters are summaries of the three most enriched gene sets from KEGG, Reactome, GO Biological Processes or GO molecular function.

### Relationship of putative acne risk genes with Mendelian and common complex skin diseases and cancer

Previous investigation of genes implicated in acne risk has revealed a striking overlap with genes that are disrupted by rare alleles in Mendelian skin disorders.^14^ Across the set of putative causal genes at the acne risk loci we note a further 41 have been observed as the site of rare alleles in Mendelian disorders (***Supplementary Table 17***). This includes *JUP, KRT16* and *RHBDF2*, rare alleles of which are responsible for Mendelian disorders in which palmoplantar keratoderma is a consistent feature (OMIM: 601214, 613000 and 148500) and *TP63* in which rare protein altering alleles cause a dominant ectodermal dysplasia. Notably, *TP53* the most frequently mutated gene in human cancers, including cutaneous melanoma,^23^ is also located at the acne susceptibility locus at 17p13.1. *MDM2* and *MDM4*, situated at acne risk loci 12q15 and 1q32.1 respectively, are potent p53 inhibitors with established roles in cellular proliferation and apoptosis whose disruption contributes to melanoma prognosis.^24^ Furthermore, the p53 regulated proto-oncogene *MYC* is also located at the acne risk locus at 8q24.21.

At 1q21.3, the allele with the strongest evidence of association with acne is a nonsense allele in the gene encoding filaggrin (*FLG*) (rs61816761, p.Arg501* OR = 0.76; 95% CI 0.69– 0.83; P = 3×10^−10^). This gene was originally described as the site of biallelic loss-of-function alleles in recessive forms of ichthyosis vulgaris, but loss of filaggrin function is now recognised as a major risk factor for atopic dermatitis.^25^ The nonsense allele which confers protection against acne has been shown to increase risk of atopic dermatitis and suggests a directionally opposing relationship between loss-of-function of filaggrin and these two highly prevalent skin conditions. Estimates of genetic correlation between acne and atopic dermatitis (rg = −0.00097; s.e. = 0.052; p = 0.83) suggest that an inverse relationship in genetic risk between these two common skin conditions is not prevalent across the genome. However, we note that the lead variants at three additional acne susceptibility loci are also genome-wide significant in the largest recent GWAS of atopic dermatitis (***Supplementary Table 18***).^25^ At two of these loci, colocalisation analysis strongly indicates that the causal variant differs between acne and atopic dermatitis, whereas at 11q22.2 colocalisation supports a shared causal variant. At this locus, the putative causal variant disrupts an enhancer sequence that is predicted to regulate the expression of *MMP3*, which encodes a matrix metalloproteinase with a role in promoting differentiation of keratinocytes during tissue remodelling. At 11q22.2 the allele that confers risk of acne is protective for atopic dermatitis, which is consistent with the direction of allelic effects observed at the filaggrin locus.

### Genetic validation of the targets of acne therapies

To evaluate whether acne associated genetic variation implicates the targets of existing or potential future acne treatments, we mapped the putative causal genes to the targets of approved therapeutic agents. Of the 243 prioritised genes, 32 are the targets for therapeutic agents. This includes genes at four acne risk loci that encode the targets for prescription acne medications or the active ingredients of widely available cosmetic products that have been demonstrated to improve acne symptoms (***Supplementary Table 19***).

These four loci include 12q24.31 where two genes (*HCAR2* and *HCAR3*) encode hydroxycarboxylic acid receptors that are activated by niacin (nicotinic acid, also known as vitamin B3), whose topical application is effective in treating acne symptoms and is the active ingredient of many cosmetic products.^5^ Similarly, *AKR1C1*, which encodes an aldo-keto reductase enzyme,at the 10p15.1 locus, is inhibited by salicylic acid and its derivatives, which are common active ingredients of cosmetic skin care formulations that promote epidermal shedding and reducing the risk of follicular occlusion.^5^ At 9q32, *ALAD* encodes delta-aminolevulinate dehydratase, also known as porphobilinogen synthase, an enzyme that acts upon aminolevulinic acid, the common precursor of all tetrapyrrole compounds, including chlorophyll, heme and vitamin B12. Topical application of aminolevulinic acid is an effective therapy for actinic keratosis and there is emerging evidence of its effectiveness for the treatment of severe acne in combination with photodynamic therapy.^5^ Whist there is no evidence of co-localisation between the acne association signal and an eQTL for *ALAD* in bulk skin, we note there is strong evidence for colocalisation in cultured fibroblasts (PP.H4 – 99.6%). The allele of the lead variant (rs1771220) in this association signal that confers protection to acne is associated with elevated expression of *ALAD* in fibroblasts.

Importantly, *RXRA*, located within the acne risk locus at 9q34.2, encodes the retinoid X receptor alpha, a nuclear receptor that is the target of topical and systemic retinoid therapies, the most widely prescribed class of acne medications.^5^ The variant with the strongest evidence of association with acne at 9q34.2, rs7852075, is located within an experimentally validated USF1 transcription factor binding site. The presence of the acne risk allele (T), which has an allele frequency of 0.61 in Europeans, at this binding site matches the USF1 consensus binding motif. This variation in the binding capability of this motif by rs7852075 is predicted, by the activity-by-contact model, to impact the transcriptional regulation of *RXRA*.

Considering genetic support for potential novel therapies for acne, it is potentially noteworthy that the gene encoding the target of PPAR-γ agonists is prioritized as a putative causal gene at 3p25.2. PPAR-γ agonists, including thiazolidinediones, are approved for the management of type 2 diabetes. Due to the critical role of this nuclear receptor in the differentiation of stem cells into sebocytes in the pilosebaceous unit,^26^ this class of drugs has been proposed as a potential therapy for acne,^27^ however no human trials have been reported to date.

### Acne susceptibility, retinoic acid signalling and the orchestration of stem cell lineage choices

All trans retinoic acid (atRA) is derived from retinol (vitamin A) and affects multiple biological processes via the nuclear retinoic acid receptor (RAR) and retinoid X receptor (RXR), which bind to specific DNA sequences as heterodimers (RXR-RAR) or homodimers (RXR-RXR) and influence transcriptional activity.^28^ Exogenous atRA and its derivatives, delivered topically or systemically, are highly effective for the treatment of acne. We sought to investigate whether genetic associations with acne impact processes upstream or downstream of atRA signalling to provide insights into the specificity of this pathway’s contribution to acne risk.

We first investigated whether variation in circulating levels of the atRA precursor, vitamin A, has a causal relationship with acne. A two sample Mendelian randomisation analysis with a genetic instrument comprising eight variants derived from a recent GWAS of circulating vitamin A levels does not support a causal relationship with acne (***Supplementary Tables 20-21***).^29^ We note that for one of the variants included in the instrument, at 7q11.23, there is evidence of a shared effect influencing both acne risk and circulating vitamin A (PP.H4 = 66.4%). There is also strong evidence that this association signal influences the expression of *MLXIPL* in skin (eQTL colocalization; PP_se_ = 0.97 and PP_NSE_ = 0.99) likely through the disruption of an enhancer sequence that has strong evidence of activity-by-contact to *MLXIPL* (***Supplementary Table 12***). Triangulation of the allelic effects across these association signals indicated that acne risk is associated with higher expression of MLXIPL and higher levels of circulating vitamin A.

To investigate genetic risk of acne downstream of atRA signalling we first investigated the enrichment of acne susceptibility alleles in experimentally derived *RXRA* transcription factor binding sites. A partitioned heritability approach (***Methods***) indicates an enrichment of acne risk at these binding sites (p = 2×10^−22^ in LoVo intestinal cell lines; ***Supplementary Table 22***). Further investigation of lead variants, and variants in high LD (r2 > 0.8), revealed disruption of *RXRA* transcription factor binding sites at 13 of the 162 acne susceptibility loci (***Supplementary table 23***). We also investigated genetic risk of acne downstream of atRA signalling by evaluating how many the putative causal genes at acne susceptibility loci are documented as direct or indirect targets of atRA.^30^ Of 532 atRA targets, 15 were prioritised as putative causal genes (enrichment OR = 2.43, p = 0.002; ***Supplementary Table 24***). This includes *SOX9*, which encodes a key effector of atRA’s recently described role in local control of lineage plasticity in the stem cell niche of the pilosebaceous unit.^31^ In addition, putative causal genes at acne susceptibility loci whose expression is affected by atRA include *WNT3A*, *TGFB2*, *GATA6* and *PPARG*, all with established roles in in cellular differentiation pathways in the skin and upper hair follicle.^31–33^

## Discussion

Over 85% of the population report having experienced acne with varying degrees of severity.^1^ From this perspective, acne can be considered a quantitative trait, where the propensity of any individual to develop acne reflects the susceptibility of each of the pilosebaceous units on the face, neck, upper chest, shoulders, and back to develop into an acneiform lesion. This conceptualisation is consistent with the notion that the cumulative risk of individual pilosebaceous units to enter into an acne cycle contributes to extent of acne symptoms.

The spectrum of acne severity in the population presents challenges in the consistency of dichotomisation of the trait in different settings. The analytical approach taken in this study aimed to address the impact of variation in dichotomisation, driven ascertainment and study design, on effect size estimates across independent GWAS of acne. The simple heuristic approach taken is unlikely to capture the full range of ascertainment differences between studies, particularly biases in healthcare-seeking behaviours and recall effects, which are likely to vary across geographical regions, age ranges, and genders. Indeed, the lead variants of 32 of the identified risk loci have nominally significant (p < 0.05) evidence of heterogeneity, indicating that there may be further sources of heterogeneity that are unaccounted for. Nevertheless, model deviance measures indicate that overall, the FIMR model fits the observed data better than the FEMA model at the majority of risk loci. Further support for the use of the FIMR approach arises from the improved calibration of effect size estimates, demonstrated by the predictive performance of a polygenic risk score (PRS) derived from this ascertainment adjusted analysis compared to a PRS derived from a FEMA. The FIMR approach also provides a rational framework for the conversion of effect estimates to the liability scale. Previous studies have demonstrated that misparameterisation of this conversion with respect to phenotype classification can lead to large upwards bias of liability scale heritability estimates.^34^ The liability-scale SNP-based heritability estimated from the FIMR of 13.4%, sits between previous estimates of 9.4% and 22.95% calculated in previous GWAS where a population prevalence of acne was assumed to be 30%.^12,15^

Investigation of genetic susceptibility to acne across ancestry groups provides evidence that the genetic architecture of acne is largely shared but supports ancestry-specific effects at some loci. The sample sizes for non-European populations are smaller and therefore power to detect effects of the magnitude observed in the European study is greatly reduced. However, the vast majority of lead variants at the identified acne susceptibility loci exhibit effect sizes that are consistent across populations with those observed in the analysis of individuals of European ancestry. The presence of ancestry-specific risk factors is supported by the identification of an allelic association with acne at rs2243130 in an African American population that is not observed in Europeans and the failure to observe an effect consistent with the previously reported effect of rs7531806 on acne risk in a Han Chinese population in neither the African American, South Asian nor European analyses. Given the sexual dimorphism of acne, it is notable that there is evidence of differential effects on acne risk between males and females at some risk loci. This includes 15q26.1, where the lead variant rs34560261 has a significantly larger effect on acne risk in males. Previous studies have mapped the causal variant at this locus to a substitution in a p63 transcription factor binding site, which significantly influences the expression of *SEMA4B* in the skin.^14^ This putative causal variant has been previously implicated in the susceptibility to a scarring form of alopecia that primarily affects females.^35^ With relevance to this effect, we implicate *TP63*, which encodes the p63 transcription factor at the newly identified acne susceptibility locus at 3q28, although there is no evidence of a sex specific effect on acne risk at this particular locus. The observed ancestry and sex differences underscore the need for larger genetic studies encompassing individuals of more diverse population ancestries to fully investigate extent of this heterogeneity and the molecular mechanisms through which these variants exert their phenotypic effects.

Across all analyses, genetic associations with genome-wide significant evidence were observed at a total of 165 genomic loci. Interrogation of putative causal genes at these loci revealed a series of genes that encode the molecular targets of the active ingredients of widely used acne therapeutics and skin care products that have been demonstrated to be effective in improving acne symptoms. Notably, this includes genetic support for the emerging use of 5-Aminolevulinic acid-based photodynamic therapy (ALA-PDT) in severe acne. The identification of an acne risk locus at 9q34.2, provides genetic validation of the gene encoding the target of atRA in acne. Exogenous administration of atRA and its derivatives, both topically and systemically, are established effective therapies for the resolution of acne symptoms,^5^ but their effectiveness is balanced with an adverse effect profile that includes initial skin irritation, redness and peeling (commonly referred to as retinisation). Systemic administration also carries risk of teratogenicity, and a possible association with longer term psychological and sexual side effects (erectile dysfunction and decreased libido) which has led to restrictions on the prescribing of retinoids in some countries.^6^

Investigation of genetic susceptibility downstream of atRA signalling provides important insights into the molecular and cellular mechanisms underlying susceptibility to acne. The prevailing view of acne pathogenesis assigns a major role to elevated or altered sebum production, which facilitates the growth of pathogenic strains of Cutibacterium acnes (C. acnes). However, our findings strongly implicate the molecular control of cell fate within the pilosebaceous unit as a critical component of acne risk. The pilosebaceous unit is a spatially constrained microenvironment where the orchestration of cell fates needs to be meticulously regulated to maintain homeostasis and reduce the risk of occlusion of the follicular ostia. There is a delicate balance between cell differentiation, proliferation, migration and apoptosis. Lineage plasticity of the stem cell niche which enables flexibility in differentiation paths based on microenvironmental cues, plays a significant role in this process. atRA has recently been shown to be a potent upstream regulator of this process,^31^ and our findings indicate that genetic variation at genomic loci where key regulators of this pathway are encoded influences an individual’s risk of acne. Not only do we observe an acne risk allele that disrupts the USF1 transcription factor binding site upstream of the gene encoding the RXR receptor but also at loci harbouring genes encoding a series of key downstream effector molecules critical for the determination of cell fates in the pilosebaceous unit. Notably this includes *SOX9*, encoding a transcription factor upregulated by atRA that influences the expression of genes critical for cell fate decisions,^36^ promoting flexibility to differentiation paths within the stem cell niche.^31^ The role of variation affecting cellular differentiation pathways is also evidenced through the identification of genetic risk loci encoding *WNT3A*, *GATA6*, *FGF1* and *PPARG*, at newly defined acne risk loci alongside *TGFB2* at the previously reported susceptibility locus at 1q41 This group of genes are all direct or indirect regulatory targets of atRA and have established roles in driving differentiation and proliferation of key cell populations in the pilosebaceous unit.^32^

Within the confines of the pilosebaceous unit, the efficient elimination of cells and cellular debris is critical for maintaining homeostasis. Acne risk loci harbour multiple genes involved in the formation of keratin intermediate filaments, desmosomes and hemidesmosomes; structure with established roles in keratinocyte shedding. The identification of *TP53*, mediators of its activity *MDM2* and *MDM4* and the p53 regulated *MYC* at acne susceptibility loci, suggests that disruption p53 mediated apoptosis may impair homeostasis in the pilosebaceous unit, contributing to acne risk. Notably, a recent study has demonstrated that the molecular switch enabling stem cells to phagocytose apoptotic cell corpses in the hair follicle is mediated by RXR-RXR activation.^37^ This interplay between p53-mediated apoptosis and RXR signalling may represent a key mechanism of genetic susceptibility. The implication of the p53 pathway in genetic susceptibility to acne also motivates further investigation of this pathway in the elevated melanoma rates across a 20 year follow up of individuals with a history of teenage acne in the Nurses’ Health Study II cohort.^38^

This comprehensive GWAS meta-analysis represents a significant advance in the understanding of acne’s genetic architecture, uncovering 117 novel susceptibility loci. The findings underscore the importance of inclusive research across diverse populations, revealing broad genetic effects shared across sex and ancestry, while also identifying loci with differential effects. The study highlights the potential for therapeutic innovation, providing genetic validation for both existing and emerging acne treatments and emphasises the importance of stem cell lineage plasticity and cellular fate control within individual pilosebaceous units as critical components of acne risk.

## Methods

### Meta-analysis and meta-regression

We combined summary statistics from 16 GWAS conducted in populations of European ancestry (study details in ***Supplementary Material and Table S1***). Prior to the meta-analysis, all variants aligned to positions on the human genome build GRCh37 (hg19) and variants with low imputation quality (INFO<0.7), minor allele frequency (MAF) < 0.01, missing values, P-values or standard errors ≤0, were excluded from the meta-analysis. Additional post-meta-analysis QC was performed by only including variants that were 1) present in at least 3 cohorts and 2) included in dbSNP (hg19, release 150), leading to a total of 8,395,919 variants in the combined sex meta-analysis, 8,415,312 in the female only and 8,411,912 in the male only meta-analysis.

We used a meta-regression approach to adjust for ascertainment effects that resulted in systematic heterogeneity of effect sizes across the cohorts in a meta-regression, the allelic effect of a variant in each GWAS study is modelled through a linear regression, with each effect weighted by its inverse variance. The fixed intercept meta-regression extends a fixed effect meta-analysis through the inclusion of covariates, or study-level variables, which can account for variations in effect sizes across studies. This method is particularly valuable for exploring how specific factors, such as the method of case ascertainment in this study, influence overall effect estimates.

We utilised simple heuristic approach to estimate the difference in the mean liability between cases and controls, which served as the study-level variable in the subsequent meta-regression. This measure was derived from the prevalence rates of acne at various severity levels, along with the estimated severity captured by each cohort’s ascertainment method (as detailed in ***Supplementary Table S1***). The prevalence rates, sourced from epidemiological studies, were defined as 50% for no acne, 50% for mild acne, 14% for moderate acne, and 2% for severe acne.^21,39–43^ The calculation of the mean liability based on the case and control ascertainment involved transforming the prevalence of the captured severities into standard normal distribution cut-points and then normalizing using the prevalence.

Differences in effect sizes between cohorts were observed using the slope from a Deming regression of the study effect sizes against a fixed effect meta-analysis estimate across 30 established acne risk loci where the previously reported lead variant or high LD proxy was available in each of the 16 studies. To validate the study level variable, we then regressed it on the Deming regression slopes using a linear regression weighted by the effective sample size in each study.

The meta-regression model partitions heterogeneity between GWAS studies into that which is correlated with ascertainment and that which is not and can be used for GWAS loci discovery.^44^ To model the effect of the ascertainment study-level variable *X_k_* on the effect size estimates for a genetic variant across *k* studies, we perform a fixed-effect meta-regression implemented using the ‘metafor’ package in R (v4.2.2). ^45^ For each *k*^th^ study, θ̂*_k_* the estimated effect size for the genetic variant, and 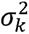 is its corresponding variance. The fixed effects meta-regression model is then formulated as:

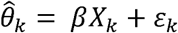

Each study’s contribution is weighted by its estimated inverse variance, 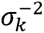. Here, we do not model an intercept term, as when *X_k_* = 0, indicating a scenario in which case/control status is randomly assigned, the expected genetic effect size *θ̂_k_* should also be zero. Also of note here, we do not estimate the between-study variance (usually denoted as *ζ_k_*, as in a random-effects meta-regression, we are therefore assuming that all the variability in the true effect sizes between cohorts is explained by the fixed parameter *βX_k_*, and any additional variation is due to sampling variance and is represented by *ε_k_* A fixed-effect meta-analysis was also performed using the ‘metafor’ package in R (v4.2.2) with the same set of input summary statistics.

### Sex stratified analysis

Sex-stratified meta-analyses across the 16 cohorts were performed to detect sex-specific genetic associations. To test for differences in effect sizes between males and females we used a post-hoc Z-difference test 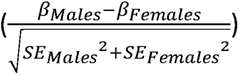. To assess statistical significance, a Bonferroni-corrected p-value threshold based on the number of loci tested was applied.

### Replication of acne susceptibility loci in South Asian and African American ancestry individuals

We assessed the replication of the acne risk loci from the current meta-analysis of European individuals in two additional cohorts, an African American cohort (570 cases and 14,134 controls) and a South Asian cohort (4094 cases and 37,935 controls). Lead SNPs identified in the European meta-analysis were assessed for significant differences in effect sizes using a post-hoc Z-difference test 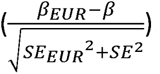.

### Susceptibility locus definition

Loci were defined as LD independent regions of the genome (Berisa & Pickrell, 2016^19^) that contain at least one SNP with P-value < 5×10^−8^. Index SNPs were defined as the SNP with the smallest P-value in LD independent regions where there is a genome-wide significant SNP. Only a single lead variant was retained in the MHC region (chr6:25-34Mb) and in the chromosome 8 inversion on 8p23.1 due to the long-range LD structure in these regions. We considered the locus to have been previously reported if it contained lead SNPs reported in published large-scale acne GWASs.^12–15^ SNPs in LD with the index SNPs were inferred based on 1000 Genomes data with European ancestry,^46^ using PLINK^47^ (version 1.90; www.cog-genomics.org/plink/1.9).

### Polygenic score analysis

The calibration of effect sizes arising from the fixed effect meta-analysis verses a meta-regression approach in which ascertainment was controlled was evaluated using polygenic risk scores in the independent PISA cohort of 2,058 individuals in which acne incidence and severity had been assessed by questionnaire (**Supplementary Note**). In this cohort acne severity was classified as none, mild, moderate and severe. PRS weights were generated using two approaches. 1. LD clumping and p-value thresholding using and series of eight p-value thresholds (5×10^−08^, 1×10^−05^, 1×10^−03^, 1×10^−02^ 5×10^−02^, 0.1, 0.5 and 1). A Bayesian regression approach, SBayesR, that estimates individual variant effects jointly with the linkage disequilibrium (LD) between them. The association between PRSs constructed from the FEMA and FIMR with acne incidence, defined as moderate or severe acne, was assessed while controlling for sex, ancestry principal components and relatedness in GCTA.

The evaluation of PRS distributions across acne severity categories and the ability to differentiate between acne these severity classifications was assessed using PRS constructed using SBayesR from the FIMR in two independent cohorts in which acne severity had been ascertained by questionnaire (PISA) and through clinical assessment of facial photographs (Generation R; **Supplementary Note**). In the self-reported cohort, acne severity was classified as none, mild, moderate and severe and in the clinically assessed cohort acne severity was classified as clear/almost clear, mild and moderate/severe.

### SNP-heritability, functional, and cell-type enrichment

LD-score regression^18^ was used to estimate the SNP-heritability (*h*^2^_SNP_) of acne based on common HapMap3^48^ SNPs and LD scores computed from the 1000 Genomes subset with European ancestry. Follow up analyses estimating the genetic correlation across traits was also performed using LDSC regression software.

We used stratified LDSC-regression to partition the SNP-heritability for acne by tissues and cell-types. Genome annotations included those curated by Finucane et al., 2015^49^ & 2018^50^: (1) a baseline model with 53 overlapping functional annotations that are not cell-type specific, including genic, regulatory, conserved and epigenomic features; (2) multi-tissue annotations of gene expression,^49,50^ generated with data from GTEx^51^ and Roadmap Epigenetics Mapping Consortium.^52^ Gene expression annotations were jointly modelled with a “control” set of all genes and with the baseline model (1).

### Functional annotation of disease associated variants

The LD independent lead variants (p < 5×10^−8^ and *r*^2^ < 0.1) and along with any variant in high LD (*r*^2^ > 0.8) at all acne risk loci, except the MHC region, were annotated using Ensembl variant effect predictor (VEP).^53^ High or moderate impact annotations were retained. The same set of variants were annotated with activity-by-contact (ABC) scores that predict disruption of enhancer-gene interactions in 131 biosamples from 74 cell types and tissues.^54,55^ We reported the highest scoring gene at each locus.

### Transcript QTL colocalisation and transcriptome wide association testing

We conducted colocalization analysis to explore the relationship between acne risk alleles and expression quantitative trait loci (eQTL) in skin tissue (both sun-exposed and not sun-exposed) using data from GTEx (v8)^51^ and Twins UK.^56^ We accessed the EBI eQTL catalogue (https://www.ebi.ac.uk/eqtl/Data_access/) to gather summary statistics for all QTLs within a 1Mb window around the index SNP in each acne associated locus that was also linked to the expression of adjacent genes (with a significance level of *P* < 1×10^−4^). Bayesian colocalization analysis was conducted for the overlapping acne and QTL variants using the ‘coloc’ package (v5.2.3)^57^ in R (v4.2.2), setting the prior probability for colocalization at 10^−5^. Evidence for colocalisation was reported if the H4 posterior probability was greater than 50% and the H3 posterior probability was less than 50% to exclude loci where there is a high probability of independent signals.

We used FUSION to conduct TWAS.^58^ Pre-computed gene expression weights for skin (both sun-exposed and not sun-exposed) were obtained from the FUSION website (http://gusevlab.org/projects/fusion/). These weights, derived from GTEx v8, were based on genes with significant heritability, incorporating all samples from GTEx v8 (with sample sizes of 517 and 605 skin not sun-exposed and skin sun-exposed. The threshold for significance in TWAS associations was set at P < 1×10^−6^, corresponding to the Bonferroni-adjusted level for the number of gene-tissue pairs (20,000 test * 2 skin tissues).

### Cross locus prioritisation of putative causal genes

We calculated the polygenic priority scores using PoPs (Polygenic Priority Score)^59^, a method that is based on the intuition that genes within risk loci are more likely to be casual when they share characteristics with genes located at other risk loci. Gene-level association statistics were calculated using MAGMA (v1.10)^60^ and gene features for 18,383 genes were gathered from a range of sources including 8,479 pathways from DEPICT^61^ (sourced from GO,^62^ KEGG,^63^ Reactome,^64^ Mouse Genome Database^65^), 8,717 protein-protein interaction features from InWeb,^66^ and 40,546 gene expression-based features from 77 datasets, predominantly single-cell expression.^59^ After a feature selection step, PoPs computes a score for each gene by regressing the selected features on the MAGMA scores. To prioritise genes for each acne locus, we selected the any gene in the top 10% of the PoPs score distribution within 500 kb in either direction of the index variant.

Gene prioritisation was performed with DEPICT.^61^ DEPICT integrates 14,462 reconstructed gene sets, derived from concordant gene expression across 77,840 microarray datasets, to assign each gene across the genome a probability for membership in each of the gene sets. This information is then used to prioritise genes across the disease-associated loci that have shared predicted biological functions and to calculate an enrichment *P*-value for each of the gene sets. As input to DEPICT, we selected clumping with a *P*-value threshold of 1×10^−5^.

### Prioritisation of putative causal genes at acne susceptibility loci

Genes were prioritised if: 1) the index variant or a variants in high LD (r^2^ > 0.8) altered the protein-coding or splice site sequence 2) there was with evidence from multiple additional sources including colocalisation and TWAS, location of index SNP (or variant in high LD r^2^ > 0.8) within an enhancer sequence linked to the genes activity by the ABC method or evidence from at least one of the cross-loci methods and 3) the nearest genes for loci where no other genes were flagged by the various prioritisation methods.

### Annotation of genes with Mendelian traits and as drug targets

For each of the prioritised putative causal genes we queried the Online Mendelian Inheritance in Man (OMIM) database for links with Mendelian disorders, and two large drug databases, ChEMBL,^67,68^ and DrugBank,^69^ for drug-target interactions. We only included drugs that are approved.

### PPIs and gene-set enrichment analysis (GSEA)

The STRING database was used to create a network of prioritized acne genes, including only protein-protein interactions (PPIs) with a confidence threshold greater than 0.5. Disconnected gene clusters containing more than four genes were isolated. For each identified cluster, up to five additional protein-protein interactions (PPIs) with non-GWAS genes were added to the network. The MCL clustering algorithm was then applied to larger connected clusters to delineate further subclusters that contain at least 4 genes located across 4 loci. Gene Set Enrichment Analysis (GSEA) was employed to functionally annotate each cluster. The top six significant gene sets from KEGG, Reactome, WikiPathways, GO biological process or GO molecular function were reported.

### Mendelian randomisation

Mendelian randomization (MR) was used in follow-up analyses to investigate the causal relationship between circulating retinol and liability to acne risk. The “TwoSampleMR” package (v0.6.4) in R (v4.3.1) was used with instrumental variables defined from variants associated with the exposure that surpass genome-wide significant levels of evidence (p < 5×10^−8^). Palindromic allele combinations were excluded to avoid strand ambiguity. The instruments alleles were harmonized between the exposure and outcome datasets. We applied several MR methods, including MR Egger regression, weighted median, inverse variance weighted (IVW), simple mode, and weighted mode. An MR instrument generated with variants surpassing a less stringent significance threshold (p < 1×10^−5^) was also tested.

### Overlap between retinol targets and acne genes

To evaluate the overlap between retinol-influenced genes and the putative causal genes, we obtained a list of retinol target genes, sourced from a review paper.^30^ A Fisher’s Exact Test was subsequently performed to evaluate the statistical significance of the observed overlap using a background set of protein coding genes from GENCODE v26 to create a 4×4 contingency table. This test calculates an odds ratio and p-value, assessing whether retinol-influenced genes are overrepresented among the acne-associated genes compared to what would be expected by chance.

We used the regulatory element locus intersection (RELI) algorithm to identify whether there was significant enrichment of transcription factor binding sites for RXRA across acne risk loci.^70^ RELI evaluates the overlap of GWAS loci (index SNPs and those in high LD with r^2^ > 0.8) and TF-bound DNA sequences determined by chromatin immunoprecipitation and sequencing (ChIP-seq). This is done by comparing the intersection counts to a null distribution that considers allele frequency and LD block structure.

Additionally, the partitioned SNP heritability of the list of retinol-influencing genes from the review paper, as well as the TF binding sites and the 100 kb regions flanking them was calculated using LDSC regression.^49,50^

## Data Availability

All data produced in the present study are available upon reasonable request to the authors

## Acknowledgements

This work was supported by an award from the Leo Foundation (LF-OC-22-001033). BLM is supported by an NMHRC Emerging Leadership Fellowship (GNT2017176). This research was funded/supported by the King’s Health Partners Centre for Translational Medicine.

We want to acknowledge the participants and investigators of the Estonian Biobank research team (Lili Milani, Reedik Mägi, Mari Nelis and Georgi Hudjashov) for data collection, genotyping, QC and imputation. This work was supported by the Estonian Research Council grants TK (TK214) and PRG1189. Computations were performed at the High Performance Computing Center, University of Tartu.

We are extremely grateful to all the families who took part in this study, the midwives for their help in recruiting them, and the whole ALSPAC team, which includes interviewers, computer and laboratory technicians, clerical workers, research scientists, volunteers, managers, receptionists and nurses.

The UK Medical Research Council and Wellcome (Grant ref: 217065/Z/19/Z) and the University of Bristol provide core support for ALSPAC. GWAS data was generated by Sample Logistics and Genotyping Facilities at Wellcome Sanger Institute and LabCorp (Laboratory Corporation of America) using support from 23andMe. This publication is the work of the authors and Josine Min will serve as guarantors for the contents of this paper.

We want to acknowledge the participants and investigators of the FinnGen study.

Genes & Health is/has recently been core-funded by Wellcome (WT102627, WT210561), the Medical Research Council (UK) (M009017, MR/X009777/1, MR/X009920/1), Higher Education Funding Council for England Catalyst, Barts Charity (845/1796), Health Data Research UK (for London substantive site), and research delivery support from the NHS National Institute for Health Research Clinical Research Network (North Thames). Genes & Health is/has recently been funded by Alnylam Pharmaceuticals, Genomics PLC; and a Life Sciences Industry Consortium of AstraZeneca PLC, Bristol-Myers Squibb Company, GlaxoSmithKline Research and Development Limited, Maze Therapeutics Inc, Merck Sharp & Dohme LLC, Novo Nordisk A/S, Pfizer Inc, Takeda Development Centre Americas Inc.

We thank Social Action for Health, Centre of The Cell, members of our Community Advisory Group, and staff who have recruited and collected data from volunteers. We thank the NIHR National Biosample Centre (UK Biocentre), the Social Genetic & Developmental Psychiatry Centre (King’s College London), Wellcome Sanger Institute, and Broad Institute for sample processing, genotyping, sequencing and variant annotation. This work uses data provided by patients and collected by the NHS as part of their care and support. This research utilised Queen Mary University of London’s Apocrita HPC facility, supported by QMUL Research-IT, http://doi.org/10.5281/zenodo.438045. We thank: Barts Health NHS Trust, NHS Clinical Commissioning Groups (City and Hackney, Waltham Forest, Tower Hamlets, Newham, Redbridge, Havering, Barking and Dagenham), East London NHS Foundation Trust, Bradford Teaching Hospitals NHS Foundation Trust, Public Health England (especially David Wyllie), Discovery Data Service/Endeavour Health Charitable Trust (especially David Stables), Voror Health Technologies Ltd (especially Sophie Don), NHS England (for what was NHS Digital) - for GDPR-compliant data sharing backed by individual written informed consent. Most of all we thank all of the volunteers participating in Genes & Health.

The Genes and Health Research Team: Eamonn Maher, Shabana Chaudhary, Joseph Gafton, Karen A Hunt, Shapna Hussain, Kamrul Islam, Hilary Martin, Mohammed Bodrul Mazid, Elizabeth Owor, Jessry Russell, Nishat Safa, John Solly, Marie Spreckley, David A Van Heel, Jan Whalley, Ishevanhu Zengeya, Emily Mantle, Shaheen Akhtar, Samina Ashraf, Dan Mason, John Wright, Daniel MacArthur, Michael Simpson, Richard C Trembath, Gerome Breen, Raymond Chung, Sang Hyuck Lee, Omar Asgar, Joanne Harvey, Karen Tricker, Caroline Winckley, Hanifa Khatun, Amna Asif, Claudia Langenberg, Grainne Colligan, Ceri Durham, Bill Newman, Ahsan Khan, Teng Heng, Matt Hurles, Vivek Iyer, Georgios Kalantzis, Vladimir Ovchinnikov, Iaroslav Popov, Klaudia Walter, Panos Deloukas, David Collier, Ana Angel, Saeed Bidi, Fabiola Eto, Sarah Finer, Chris Griffiths, Sam Hodgson, Benjamin M Jacobs, Rohini Mathur, Caroline Morton, Asma Qureshi, Stuart Rison, Annum Salman, Miriam Samuel, Moneeza K Siddiqui, Daniel Stow, Sabina Yasmin, Julia Zöllner, Sheik Dowlut

The Lifelines initiative has been made possible by subsidy from the Dutch Ministry of Health, Welfare and Sport, the Dutch Ministry of Economic Affairs, the University Medical Center Groningen (UMCG), Groningen University and the Provinces in the North of the Netherlands (Drenthe, Friesland, Groningen).

The Trøndelag Health Study (The HUNT Study) is a collaboration between HUNT Research Centre (Faculty of Medicine and Health Sciences, NTNU, Norwegian University of Science and Technology), Trøndelag County Council, Central Norway Regional Health Authority, and the Norwegian Institute of Public Health. LT, KH and ML work in a research unit funded by Faculty of Medicine and Health Sciences, NTNU, Norwegian University of Science and Technology; The Liaison Committee for education, research and innovation in Central Norway; the Joint Research Committee between St. Olavs Hospital (Trondheim, Norway) and the Faculty of Medicine and Health Sciences, NTNU, Norwegian University of Science and Technology. ML is supported by grants from the Liaison Committee for education, research and innovation in Central Norway. The genotyping in HUNT was financed by the National Institutes of Health; University of Michigan; the Research Council of Norway; the Liaison Committee for Education, Research and Innovation in Central Norway; and the Joint Research Committee between St Olavs hospital and the Faculty of Medicine and Health Sciences, NTNU.

We thank the funders and contributors to the previous two King’s College London led acne GWAS studies, Petridis et al. (2018): Christos Petridis, Alexander A. Navarini, David Baudry, Michael Duckworth, Michael H. Allen, Charles J. Curtis, Sang Hyuck Lee, A. David A. Burden, Alison Layton, Veronique Bataille, Andrew E. Pink, The Acne Genetic Study Group, Isabelle Carlavan, Johannes J. Voegel, Timothy D. Spector, Richard C. Trembath and John A. McGrath; and Navarini et al. (2014): Michael Weale, Jo Knight, Pascale Reiniche, Robert Pleass, Daniel Creamer, John English, Stephanie Munn, Shernaz Walton, Carolyn Willis and Sophie Déret. We thank Alka Saxena; Ulrike Blume-Peytavi; Leaca Crawford; Jana Estafan; Darren Geoghegan; Dan Glass; Alison Gosh; Naomi Hare; Helen Holmes; Karen Markwell; Eleanor Mallon; Philippe Martel; Carine Marty; Corinne Ménigot; Efterpi Papouli; Anne Thompson; Kate Thornberry; Bianca Tobin, and the participating patients and supporting staff in all the study centres.

Support was provided by the National Institute for Health Research (NIHR), through the Dermatology Clinical Research Network; the NIHR Biomedical Research Centre based at Guy’s and St Thomas’ NHS Foundation Trust and King’s College London; Galderma, Bruno Bloch, Promedica Foundation and HSM-2 canton of Zurich, British Skin Foundation; Medical Research Council/British Association of Dermatologists/British Skin Foundation Clinical Research Training Fellowship; the NIHR Maudsley Biomedical Research Centre at South London and Maudsley NHS Foundation Trust and King’s College London; capital equipment funding from the Maudsley Charity (Grant Ref. 980) and Guy’s and St Thomas’ Charity (Grant Ref. STR130505).

We want to acknowledge Partners HealthCare Biobank for providing samples, genomic data, and health information data. XD is supported by American Parkinson’s Disease Association, and NIH U01.

We thank all twins and family members for their participation. Analyses were supported by the Netherlands Organization for Scientific Research: Netherlands Twin Registry Repository: researching the interplay between genome and environment (480-15-001/674); the European Union Seventh Framework Program (FP7/2007-2013): ACTION Consortium (Aggression in Children: Unravelling gene-environment interplay to inform Treatment and InterventiON strategies; grant number 602768). Genotyping was made possible by grants from NWO/SPI 56-464-14192, Genetic Association Information Network (GAIN) of the Foundation for the National Institutes of Health, Rutgers University Cell and DNA Repository (NIMH U24 MH068457-06), the Avera Institute, Sioux Falls (USA) and the National Institutes of Health (NIH R01 HD042157-01A1, MH081802, Grand Opportunity grants 1RC2 MH089951 and 1RC2 MH089995) and European Research Council (ERC-230374). DIB acknowledges her KNAW Academy Professor Award (PAH/6635). MB is supported by an ERC consolidator grant (WELL-BEING 771057 PI Bartels) and NWO VICI grant (VI.C.2111.054, PI Bartels).

We are indebted to all participants for giving their time to contribute to these studies. Additionally, we wish to thank all the people who helped in the conception, implementation, media campaign and data cleaning. In particular, we would like to thank our research nurses - including Ann Eldridge and Marleen Grace, who were responsible for collecting the acne data in our young twin cohort under the supervision of Dr. Margie Wright. We would like to thank Scott Gordon who curated all the genotyping data that was used in this project. Data collection for the Australian Genetics of Depression Study (self-report cohort) was possible, thanks to funding from the National Health and Medical Research Council (NHMRC) of Australia grant 1086683. This work was further supported by NHMRC grants 1145645, 1078901 and 1087889. PISA is funded by a NHMRC Boosting Dementia Research Initiative grant, APP1095227, to NGM and others. MKL is supported by a Boosting Dementia Leadership Fellowship (APP1140441). MER thanks support of the NHMRC and Australian Research Council (GNT1102821). BLM was supported by an NHMRC EL1 fellowship (2017176)

This research has been conducted using the UK Biobank Resource (approved project 15147). It uses data provided by patients and collected by the National Health Service (NHS) as part of their care and support. Copyright 2024, NHS England. N.D. is funded by Health Data Research UK (MR/S003126/1), which is funded by the UK Medical Research Council, Engineering and Physical Sciences Research Council; Economic and Social Research Council; Department of Health & Social Care (England); Chief Scientist Office of the Scottish Government Health and Social Care Directorates; Health and Social Care Research and Development Division (Welsh Government); Public Health Agency (Northern Ireland); British Heart Foundation; and Wellcome.

The QSkin study was funded by the NHMRC (grant numbers 1073898, 1063061 and 1185416). DCW is supported by a NHMRC Investigator Grant (2026567).

We are grateful to all study participants for their support of and contributions to this study. This phase of the eMERGE Network was initiated and funded by the NHGRI through the following grants: U01HG008657 (Group Health Cooperative/University of Washington); U01HG008680 (Columbia University Health Sciences); U01HG008685 (Brigham and Women’s Hospital); U01HG008672 (Vanderbilt University Medical Center); U01HG008666 (Cincinnati Children’s Hospital Medical Center); U01HG006379 (Mayo Clinic); U01HG008679 (Geisinger Clinic); U01HG008684 (Children’s Hospital of Philadelphia); U01HG008673 (Northwestern University); U01HG008701 (Vanderbilt University Medical Center serving as the Coordinating Center); U01HG008676 (Partners Healthcare/Broad Institute); U01HG008664 (Baylor College of Medicine); and U54MD007593 (Meharry Medical College). NCATS TL1TR001875 (to A.K.), NIAMS K01AR075111 (to L.P.) and NIAMS R01AR080796 (to L.P).

The Generation R Study is conducted by the Erasmus Medical Center in close collaboration with the Erasmus University Rotterdam, School of Law and Faculty of Social Sciences, the Municipal Health Service Rotterdam area, the Rotterdam Homecare Foundation, and the Stichting Trombosedienst and Artsenlaboratorium Rijnmond (STAR). The authors gratefully acknowledge the contribution of general practitioners, hospitals, midwives, and pharmacies in Rotterdam. Financial support for the Generation R Study comes from the Erasmus Medical Center, Rotterdam, Erasmus University Rotterdam, and the Netherlands Organization for Health Research and Development (ZonMw).

## Notes

### Competing Interest Statement

The authors have declared no competing interest.

### Summary of Updates

Author list and affiliations corrected.

