## Supplemental note for "Genome-wide association meta-regression identifies stem cell lineage orchestration as a key driver of acne risk" (from another version of this preprint: version 1, 2025-06-28)

1. **SUPPLEMENTARY NOTE: ACNE COHORT DETAILS**

**1. ALSPAC**

ALSPAC (The Avon Longitudinal Study of Parents and Children) is a transgenerational prospective observational study investigating influences on health and development across the life course. Pregnant women resident in Avon, UK with expected dates of delivery 1st April 1991 to 31st December were invited to take part in the study. Of the 14,541 enrolled pregnancies, there were a total of 14,676 foetuses, resulting in 14,062 live births and 13,988 children who were alive at 1 year of age.^1,2^ When the oldest children were approximately 7 years of age, an additional 913 pregnancies were enrolled resulting in a total sample size of 15,589 foetuses. Of these 14,901 were alive at 1 year of age. Participants attended clinics at the University of Bristol and filled out follow-up questionnaires at various intervals. Please note that the study website contains details of all the data that is available through a fully searchable data dictionary and variable search tool" and reference the following webpage: http://www.bristol.ac.uk/alspac/researchers/our-data/

Ethical approval for the study was obtained from the ALSPAC Ethics and Law Committee and the Local Research Ethics Committees. Consent for biological samples has been collected in accordance with the Human Tissue Act (2004). Informed consent for the use of data collected via questionnaires and clinics was obtained from participants following the recommendations of the ALSPAC Ethics and Law Committee at the time.

Children were genotyped using the Illumina HumanHap550 quad genome-wide SNP genotyping platform (Illumina Inc., San Diego, CA, USA) by the Wellcome Trust Sanger Institute (WTSI; Cambridge, UK) and the Laboratory Corporation of America (LCA, Burlington, NC, USA). Individuals were excluded on the basis of incorrect gender assignment, abnormal heterozygosity (<0.320 or >0.345 for WTSI data; <0.310 or >0.330 for LCA data), high missingness (>3 %), cryptic relatedness (>10 % identity by descent) and non-European ancestry (detected by MDS analysis). Following QC, the final directly genotyped dataset contained 500,527 SNP loci. Mothers were genotyped using the Illumina Human660W-quad genome-wide SNP genotyping platform (Illumina Inc., San Diego, CA, USA) at the Centre National de Génotypage (CNG; Paris, France). Individuals were excluded based on non-European ancestry, missingness, relatedness, gender mismatches and heterozygosity. PLINK (v1.07) was used to carry out QC measures on an initial set of 10,015 subjects and 557,124 directly genotyped SNPs. Following QC, the final directly genotyped dataset contained 526,688 SNP loci. The ALSPAC genotype data generated using the Illumina HumanHap550 quad (children) and Illumina human660W quad (mothers) has been jointly phased using SHAPEIT v2 (which uses relationship information to improve phasing accuracy) and imputed to the HRC panel (HRC.r1.1)^3^ using the Michigan imputation server.^4^

Acne data was collected via clinical assessment at various time-points. Here, the 13.5 years old time-point was selected as a compromise between the number of individuals with data at the time-point (at each time-point, there was attrition of patients) and having an older age to allow for the maximum number of acne cases that manifest during puberty. If any acne variants were present on any of the three main sites (face, chest, back) the severity was recorded according to the “Acne Grading Guide” as trivial, mild, moderate or severe.^5^ There were 5,069 participants with GWA data and acne data at age 13.5 years. Acne cases were defined as individuals with “severe” and “moderate” acne (193) and controls as “trivial” and “no acne” (3,772). GWAS was run in SNPTESTv2.5.2 using age, sex and 20 ancestry principal components as covariates.

**2. FINNGEN**

FinnGen data consists of 377,277 Finnish individuals (166,407 males and 210870 females), which includes prospective epidemiological and disease-based cohorts as well as hospital biobank samples (listed at <https://finngen.gitbook.io/documentation/>). The data were linked by the unique national personal identification numbers to national hospital discharge, death, and medication reimbursement registries. The Ethical Review Board of the Hospital District of Helsinki and Uusimaa approved the FinnGen study protocol (HUS/990/2017). Participation in FinnGen utilizing biobank samples is always voluntary and sample donors are free to cancel, change or restrict their consent to biobank research at any time.

Chip genotype data processing and QC Samples were genotyped with Illumina (Illumina Inc., San Diego, CA, USA) and Affymetrix arrays (Thermo Fisher Scientific, Santa Clara, CA, USA). Genotype calls were made with GenCall and zCall algorithms for Illumina and AxiomGT1 algorithm for Affymetrix data. Chip genotyping data produced with previous chip platforms and reference genome builds were lifted over to build version 38 (GRCh38/hg38) following the protocol described here: dx.doi.org/10.17504/protocols.io.nqtddwn. In sample-wise quality control, individuals with ambiguous gender, high genotype missingness (>5%), excess heterozygosity (+-4SD) and non-Finnish ancestry were excluded. In variant-wise quality control variants with high missingness (>2%), low HWE P-value (<1e-6) and minor allele count, MAC<3 were excluded. Prior to imputation, chip genotyped samples were pre-phased with Eagle 2.3.5 (<https://data.broadinstitute.org/alkesgroup/Eagle/>) with the default parameters, except the number of conditioning haplotypes was set to 20,000. Genotype imputation was done with the population-specific SISu v3 reference panel. A variant call set was produced with the GATK HaplotypeCaller algorithm by following GATK best practices for variant calling. Genotype-, sample- and variant-wise QC was applied in an iterative manner by using the Hail framework v0.1, and the resulting high-quality WGS data for 3,775 individuals were phased with Eagle 2.3.5 as described in the previous section. Genotype imputation was carried out by using the population-specific SISu v3 imputation reference panel with Beagle 4.1 (version 08Jun17.d8b). Post-imputation quality-control involved checking expected conformity of the imputation INFO-value distribution, MAF differences between the target dataset and the imputation reference panel and checking chromosomal continuity of the imputed genotype calls.

Data Finnish Care Register for Health Care (CRHC) database maintained by the Finnish Institute of Health and Welfare.were used to define disease endpoints, harmonising over ICD revisions 8, 9 and 10. In the CRHC, in-patient visits are recorded from 1987 and outpatient visits from 1998 onwards. The Finnish Care Register for Health Care contains data on patients discharged from inpatient care, day surgeries and specialized outpatient care. Data contains information e.g. about providers of hospital services and specialized health care, patients, admission and discharge, diagnoses, and treatments provided. It does not include data about behavioral matters (e.g. smoking) or about socioeconomical status. Detailed description about data contents can be found in [https://www.thl.fi/en/web/thlfi-en/statistics/information-on-statistics/register-descriptions/care-register-for-health-care](https://eur03.safelinks.protection.outlook.com/?url=https%3A%2F%2Fwww.thl.fi%2Fen%2Fweb%2Fthlfi-en%2Fstatistics%2Finformation-on-statistics%2Fregister-descriptions%2Fcare-register-for-health-care&data=05%7C01%7Cmichael.simpson%40kcl.ac.uk%7C2e36eaf128024538a04308dab0db8bfc%7C8370cf1416f34c16b83c724071654356%7C0%7C0%7C638016754274097089%7CUnknown%7CTWFpbGZsb3d8eyJWIjoiMC4wLjAwMDAiLCJQIjoiV2luMzIiLCJBTiI6Ik1haWwiLCJXVCI6Mn0%3D%7C3000%7C%7C%7C&sdata=Gq16zJGPJm2jc1llNJqUeqwhKpQawLF5h6NtcOLzURY%3D&reserved=0). We selected the acne endpoint (ICD-10: L70), which had 13,981 cases (4567 males and 9414 females). This endpoint excluded any individual with ICD-10 codes L60-75 (Disorders of skin appendages) from controls, leaving 363,296 individuals (161,840 males and 201456 females).

Regenie software (version 3.2.2)^6^ was used for running the GWAS. The model included sex (in the combined male+female GWAS only), age, 10 PCs and genotyping batch as covariates. Downloaded FinnGen summary statistics were lifted over from build hg38 to hg19 using the UCSC liftover tool (downloaded 11 June 2020; <http://genome.ucsc.edu>).

**3. LIFELINES**

LifeLines is a population-based cohort study and biobank, in which participants and their family members were enrolled during the period 2003 to 2013.^7^ In all 167,729 participants were enrolled and screened at baseline. Results from extensive questionnaires and data generated from physical examinations are available. Participants were invited for re-examination in 5-year intervals and were asked to fill out further questionnaires in intervening periods.

At the time of writing, 72.433 LifeLines participants have been genotyped, all analyses that we report have been carried out on these individuals. Genotyping was carried out using three SNP chips, and results were separately imputed:

- GSA: Infinium Global Screening Array (GSA), imputed using HRC panel 1.1.^3^ as implemented by the Sanger Imputation Service at the time (24-09-2019). Imputation was done based on the autosomes (569660 SNPs that passed all QC filters) for 36,339 participants in UGLI.
- CytoSNP: HumanCytoSNP-12v2 and HumanCytoSNP- 12v2.1. Imputation was performed using IMPUTE2 (version 2.3.2), using the combined reference panel of GoNL and 1000G.
- Affymetrix: FinnGen Thermo Fisher Axiom® custom array. A final set of 28,250 samples and 462,731 markers on autosomal and X chromosomes passing all QC steps described above were used for genetic imputation. Genetic imputation was done through the Sanger imputation service using the Haplotype Reference Consortium (<http://www.haplotype-referenceconsortium.org>) panel.

All genotype imputation was carried out using genome build 37. CytoSNP samples with duplicates or close relatives in either other chipset were removed to avoid sample overlap; as were GSA samples with duplicates or close relatives in the Affymetrix chipset. The CytoSNP chip is the lowest quality of the three, so these samples were prioritized for removal.

Questionnaire data relevant to acne is available at various times (at baseline, at first follow-up, second follow-up, etc.). Each questionnaire contains different questions relevant to the presence of acne. For the child questionnaires these include questions regarding the use of acne medication. The questionnaires for children and adults were different, note that the adult questionnaire does not contain questions regarding medication use.

A phenotype “acne ever” was defined in the following way. For adults, the variable was set to 1 if any of the following acne-related questions were answered as currently having, or ever having had, acne; and 0 otherwise:

- acne presence adu q 1: the presence of acne in baseline questionnaire (encoded as 1 if yes; missing if no).
- acne presence adu q 2: the presence of acne in the second questionnaire(encoded as 0 (no), 1 (yes, now), 2 (yes, in the past).)

For children, the variable was set to 1 if any of the following questions was answered as currently having, or ever having had, acne of any severity; and 0 otherwise:

- acne severity chi q 1, (ordinal encoding, with 0 = no acne, . . . , 4 = severe acne)
- acne medication ch3a q 1, (binary encoding)
- acne medication ch3 q 1, (binary encoding)
- acne medication ch4 q 1, (binary encoding)
- acne presence ch1 q 1, (binary encoding)
- acne presence ch3a q 1, (binary encoding)
- acne presence ch3 q 1, (binary encoding)
- acne presence ch4 q 1, (binary encoding)
- acne treatment ch3a q 1, (binary encoding)
- acne treatment ch3 q 1, (binary encoding)
- acne treatment ch4 q 1, (binary encoding)

This phenotype definition results in the following case numbers:

|  |  |  | Cases | Controls |
| --- | --- | --- | --- | --- |
| GSA | Males  Females | 11958  16711 | 385  609 | 11573  16102 |
|  | Total | 28785 | 1002 | 27783 |
| CytoSNP | Males  Females | 6431  8958 | 214  324 | 6217  8634 |
|  | Total | 15399 | 538 | 14861 |
| Affy | Males  Females | 10703  17538 | 349  629 | 10354  16909 |
|  | Total | 28249 | 978 | 27271 |

A list of samples using PLINK (PLINK/1.9-beta6-20190617)^8^ was created and subsequently we used REGENIE (regenie/v3.2.2)^6^ to perform GWAS. Phenotypes were extracted from the lifelines data using Python (v3.x from Anaconda3/2022.05). Preprocessing of the affymetrix data and creation of the phenotype files was done using R (version R/4.2.2-foss-2022a-bare). To avoid batch effects, we conducted separate GWAS for the subcohorts typed on each of the three genotyping chips. In addition, we ran male-only, female-only, and combined analyses (a total of 9 analyses). We included 6 principal components to account for population stratification. Additionally, we included age as a covariate, and we used sex as a covariate in the combined (male+female) analyses.

**4. HUNT**

The Trøndelag Health Study (HUNT) is a population-based cohort study carried out at four time points over approximately 40 years (HUNT1 [1984-1986], HUNT2 [1995-1997] and HUNT3 [2006-2008] and HUNT4 [2017-2019]).^9^ All inhabitants aged 20 years and over residing in Trøndelag County in Norway were invited to participate. The surveys included clinical measurements, blood sampling and questionnaires on general health measures and a broad range of self-reported diseases and symptoms.^10^ Participants were also linked to regional- and national health registries through a unique national identification number, which enabled acne cases to be identified by searching for acne ICD codes. Participation in the HUNT Study is based on informed consent, and the study has been approved by the Data Inspectorate and the Regional Ethics Committee for Medical Research in Norway (Reference Number 27420).

Participants from HUNT2 and HUNT3 were genotyped using one of three different Illumina HumanCoreExome arrays (HumanCoreExome12 v1.0, HumanCoreExome12 v1.1 and UM HUNT Biobank v1.0). Genotype calling was performed with GenTrain v.2.0 in GenomeStudio v.2011.1 (Illumina). Samples with <99% genotype calls, with large chromosomal copy number variants, contamination >2.5% as estimated with BAF Regress,^11^ with genotypic and phenotypic sex discordance, and not of European ancestry were excluded, leaving 69,422 genotyped subjects. Genetic variants out of Hardy-Weinberg equilibrium (p-value <0.0001) were excluded.

Imputation was performed on samples of recent European ancestry using Minimac4 (v1.0.2, https://genome.sph.umich.edu/wiki/Minimac4) from a merged reference panel constructed from the Haplotype Reference Consortium (HRC) panel (v1.1) and a local reference panel based on 2,202 whole-genome sequenced HUNT participants, resulting in 24.9 million SNPs (R^2^>=0.3). Imputation on the 1000 genomes project was done using Minimac4 (v1.0.2), resulting in 15.1 million SNPs (R^2^>=0.3).

The following ICD-10 codes were used to define acne cases: L70.0 (acne vulgaris), L70.1 (acne conglobata), L70.8 (other acne), L70.9 (acne, unspecified). The ICD-9 code 706.1 was also used as it was equivalent to L70.0, L70.1 and L70.8. There were 456 individuals in HUNT with at least one of the specified codes, with the remaining 68,590 individuals defined as controls. GWAS was run in SAIGE v0.29,^12^ using sex, birth year, genotyping batch and 4 ancestry principal components as covariates. Variants with MAF >1% were included in the analyses, and dosages were used for imputed variants.

**5. King’s College London (KCL)**

The KCL datasets are two case-control study consisting of a total of 5,602 severe acne cases recruited through a network of hospital-based dermatologists within the United Kingdom and 21,120 unselected population controls. The study was designed in accordance with the Declarations of Helsinki, and ethical approval was obtained from the NRES Committee London-Westminster (reference CLRN 05/Q0702/114). The first round consisting of 1,779 acne cases and 4,976 controls from the UK population^13^ and the second round consisting of 3,823 cases and 16,144 unselected population controls. Each participant provided informed consent.

The diagnostic criteria remained the same for both rounds of recruitment, with one or more of the following criteria required for diagnosis: (a) nodulocystic disease; (b) ≥5 points in any body region assessed by the validated Leeds clinical acne score that uses a colour photographic acne grading scheme to evaluate the severity of involvement of body regions (face 0–12, chest 0–8 and back 0–8); (c) requiring treatment with isotretinoin; and (d) presence of rare and severe forms of acne.

Genotyping, QC and imputation is described in Navarini et al.^13^ and Petridis et al.^14^ for the respective studies. For both studies association testing was performed with a logistic Wald association test (EPACTS), including the first four ancestry principal components, sex and QC/imputation batch as covariates. Following this, a standard error-weighted meta-analysis was performed across the two datasets.

**6. MASS GENERAL BRIGHAM BIOBANK (PARTNERS BIOBANK)**

Samples, genomic data, and health information were obtained from the Mass General Brigham Biobank, a samples biorepository of consented patients at Mass General Brigham (parent organization of Massachusetts General Hospital and Brigham and Women’s Hospital). Data for over 35,000 volunteers is available (with informed consent).^15^ The Partners Human Research Committee (the Institutional Review Board) approved that all procedures meet ethical standards for human subject research. Participants have been genotyped using the Illumina Multi-Ethnic Global Array (MEGA) (Illumina, Inc., San Diego, CA) GWAS array. Genetic variants with a high missingness or extreme allele frequencies were removed prior to imputation using the HRCr1.1 reference panel on the Michigan Imputation server. Imputed genotype data in dosage format was used for the analysis. Acne cases were defined based on a diagnosis available on electronic health records (ICD 10 code: L70). Controls were participants not showing an acne ICD code. Participants were excluded if deemed ancestry outliers from the European population or showed high rates of missingness. GWAS were performed in PLINK v2.00 using a logistic regression adjusting for age, sex, ten genetic principal components and genotype batch as covariates.

**7. NETHERLANDS TWIN REGISTER**

The Netherlands Twin Register (NTR) is a population-based cohort that includes twins / multiples and their family members from the Netherlands.^16^ Phenotype data on acne were collected in multiple surveys. Buccal cells or blood sample for DNA isolation were collected in multiple NTR projects.^17,18^ See also: http://www.tweelingenregister.vu.nl/ for more information. All participants gave written informed consent. Studies were approved by the Central Ethics Committee on Research Involving Human Subjects of the VU University Medical Centre, Amsterdam, an Institutional Review Board certified by the U.S. Office of Human Research Protections (IRB number IRB00002991 under Federal-wide Assurance- FWA00017598; IRB/institute codes, NTR 03-180).

**7.1 Teenage cohort**

The adolescent twins and their family members included in this study were asked to report on their acne when the twins were 14, 16 and 18 years of age. Acne was ascertained by the question “Do you have pimples?” with three answer categories (“no”, “yes, a little”, and “yes, a lot”). Self-reported acne and genotypes were available for 1,819 adolescents and family members at age 14, 2,021 participants at age 16, and 688 participants at age 18. Here, participants were included if self-reported acne and genotyping were available at age 16. Otherwise, the acne measurement of age 14 or 18, depending on availability, was included. Overall, this cross-age dataset included self-reported acne and genotypes of 3,017 individuals from 1,530 families (age range: 10.5 – 35.3 years, mean age = 17.0, SD age = 1.7, 57.2% female). In total, the cross-age dataset included the acne measurements of 2021 individuals (67.0%) as measured when the twins were 16 years of age, of 493 individuals (16.3%) when the twins were 14 years of age, and of 503 individuals (16.7%) when the twins were 18 years of age. The 3-point acne scale was dichotomized by combining the “no” and “yes, a little” answer categories).

**7.2 Adult cohort**

The adult twins and their family members included in this study completed self-report questionnaires in 2000. Here, acne was ascertained by the question “Do you have pimples?” with two answer categories (“no” [i.e., ‘control’], and “yes” [i.e., ‘case’])^17^. Self-reported acne and genotypes were available for 3,091 individuals from 1,730 families (age range: 12.0 – 79.0 years, mean age = 30.9, SD age = 11.4, 67.7% females).

**7.3 Genotyping and Imputation**

For all participants, genotyping was done on several platforms, namely Perlegen Affymetrix, Affymetrix 6.0, Affymetrix Axiom, Illumina Human Quad Bead 660, Illumina Omni 1M, Illumina GSA. Part of the sample was sequenced in the Genome of the Netherlands (GoNL) project.^19^ Genotyping of each platform was performed according to the manufacturer’s protocols with the calling software as was appropriate at the time. The Single Nucleotide Polymorphisms (SNPs) of the Perlegen-Affymetrix, Illumina Human Quad Bead 660 and the Illumina Omni 1M arrays were typed on older genome builds. Therefore, these were lifted to Build 37 HG19 based on their RSid location in DBSNP 142. Within each platform and on the combined dataset, samples and SNP quality control (QC) was performed. Samples were removed if sex did not match the expected, heterozygosity F statistic was < -0.10 or > 0.10, call rate was < 0.90. SNPs were removed if minor allele frequency (MAF) was <0.01, Hardy-Weinberg Equilibrium (HWE) *P*-value < 1×10^-05^, call rate < 0.95, number of Mendel errors < 20.^8^ Furthermore, palindromic AT/GC SNPs with MAF between 0.4 and 0.5 were removed to avoid wrong strand alignment to the reference panel. The data of each platform was aligned to the GoNL reference set (V4), and SNPs were removed if they had a difference in allele frequency > 0.10, mismatching alleles, location or strand.

The data of the six platforms were combined into one dataset retaining all QCed SNPs of each platform. For individuals measured on multiple platforms, one platform was chosen in the following order: Affymetrix Axiom, Affymetrix 6.0, Illumina Omni 1M, Illumina Human Quad Bead 660, Illumina GSA, Perlegen-Affymetrix. For the SNPs included in this combined platform, using the PLINK and KING programs, DNA Identity By Descent (IBD) state was estimated for all individual pairs. The IBD estimates were compared to the expected familial relations, and samples were removed if these failed to fit. A similar approach was used for DNA zygosity mismatches. Prior to imputation, duplicate monozygotic twins (N=3,032), triplets (N=7) and NTR samples from the GONL data (N=364) were removed from the data. Then data were cross-platform imputed using MaCH-Admix against GoNL reference set to predict the missing SNPs in each platform based on haplotypes estimated from Dutch reference panel.^20^ After imputation, the GONL samples were re-added to the data, and the second (and third) MZ twin was re-added by duplicating the data of the MZ co-twin included in the imputation. SNPs were removed from the cross-platform imputed dataset if the HWE *P*-value was < 1×10^-05^, Mendel error rate was more than mean + 3SD, R^2^ imputation quality metric was < 0.90, if *P*-value for association with a platform was < 1×10^-05^.

Ancestry outliers (non-Dutch ancestry) were defined based on Principal Components Analysis (PCA) by projecting 10 Principal Components (PCs) from 1000G reference set populations on NTR cross-platform imputed data using SMARTPCA. Individuals with PC values located outside of the range of European and/or British populations were defined as outliers. Upon exclusion of outliers, 10 PCs were recomputed for NTR cross-platform imputed data to capture the variation within the Netherlands.^21^

Genotype imputation to the HRC 1.1 reference panel was done on the Michigan Imputation server on the cross-platform imputed data.^4^ The data were aligned using the PERL based "HRC Imputation preparation and checking" tool (v4.2.5; <https://www.well.ox.ac.uk/~wrayner/tools/>). The remaining SNPs (1,307,940) were phased with EAGLE for the autosomes and SHAPEIT for chromosome X before imputation using Minimach 3 following the standard imputation procedures of the server.^22^

**7.4 GWAS**

Genome-wide association (GWA) studies were performed with Scalable and Accurate Implementation of Generalized mixed models (SAIGE) v 0.39 to control for unbalanced case-control ratios and account for sample relatedness by the inclusion of a genetic relatedness matrix (GRM;^12^). The NTR cross-platform imputed dataset was used for the calculation of the GRM, and the HRC imputed data (autosomes only) was used for the single-variant association tests. The single-variant association tests employed saddle point approximation to account for case-control imbalance. In the single-variant association test, the HRC-imputed genotype data was filtered on MAF > 0.0001, Minor Allele Count (MAC) > 1 and R^2^ imputation quality metric > 0.60. Covariates included age, sex, genotyping platform and 10 ancestry PCs.

**8. QIMR BERGHOFER**

**8.1 Self Report Cohort**

Phenotypic data for acne prevalence was obtained through self-reported questionnaires in the Australian genetics of depression study (AGDS) which comprises ~20,000 participants, of which more than 18,000 have been genotyped.^23^ Briefly, the item to ascertain acne was: “*Many people suffer from acne during their lives. How much acne did you have when you were a teenager?*” Participants that responded *Never* were coded as controls, while participants that responded *Moderate* or *Severe* were coded as cases (mild cases excluded). Genotyping was conducted using the Illumina Infinium Global Screening Array platform. Prior to imputation, a common set of high QC markers between the different genotyping batches was obtained. Marker exclusion criteria included: unknown or ambiguous map position and strand alignment in a BLAST search, missingness >5%, p(HWE test)< 10^-6), MAF<1%, GenTrain score <0.6. The Michigan imputation server was used to impute the genotypes using the HRCr1.1 as a reference panel. Individuals were excluded based on high missingness (missing rate > 3%), inconsistent (and unresolvable) sex, or if deemed ancestry outliers from the European population (6 sd deviations from the first two genetic principal components from 1000 Genomes). Imputed genotype dosages were used for the analyses. GWAS was carried out in SAIGE (v0.36.3.3) in R 3.6.1 using a generalised linear mixed model to account for population stratification, cryptic relatedness and unobserved genetic confounding. The GWAS was further adjusted for age, sex, genotyping batch and the first ten principal components.

**8.2 Clinical Cohort**

Data for this cohort was collected through nurse-rated of acne for 3,817 twins and 674 siblings.^24^ The severity of acne was rated by a nurse using a 4-point scale (1 = absent; 2 = mild; 3 = moderate; 4 = severe) on back, face, and chest. Participants were part of the Brisbane Longitudinal Twin Study (BLTS) conducted at the QIMR Berghofer Medical Research Institute. Within the BLTS study, data is collected on a wide range of biological, psychological, and social traits, as well as environmental exposures. Further details of the recruitment process, data collection, and determination of zygosity can be found in Wright et al.^25^. The measures were taken longitudinally at ages 12 and 14 years and from the face only at age 16. For this analysis, we chose the score that corresponds to the most severe across all sites at all ages to minimize missing data and to allow for individual differences in the time onset of acne; the mean age of the twins was 14.6 (range 12–23, SD 1.4). The individuals in this study were genotyped on the HumanCoreExome-12v1-0_C or IlluminaHuman610WQuad bead chip. Genotypes were imputed to the HRCr1.1 reference panel. Cases were defined as having moderate or severe acne, and controls were defined as having no acne on either the back, face and chest. Mild acne cases were excluded from the analysis. GWAS was carried out in SAIGE as described above. The GWAS was further adjusted for age, sex, genotyping batch and the first ten principal components.

**8.3 Independent Sample for PRS Prediction**

Self-reported acne diagnosis from the Prospective Imaging of Aging Study (PISA) at QIMR Berghofer was used as our target sample for PRS prediction. PISA is a prospective cohort of healthy Australians at mid to late adulthood. The PISA ‘online’ component currently consists of approximately 3,000 genotyped individuals who have completed extensive behavioural, psychological and medical questionnaires, cognitive testing and linkage to medical records.^26^ Participants were asked, *“How much acne did you have when you were a teenager?”* to which they could answer *‘None’, ‘Mild’, ‘Moderate’ or ‘Severe’.* Only individuals that answered ‘Moderate’ or ‘Severe’ were included as cases in our analysis (N=527), and individuals that reported not having acne were considered controls (N=645). Genotyping was previously performed within the recruitment pool as detailed.^26,27^

All participants in all QIMR cohorts gave informed written consent; ethical approval was obtained from the QIMR Berghofer Research Ethics Committee.

**9. UK BIOBANK**

This project used data from the UK Biobank under approved project number 15147. UK Biobank is a prospective study with over 500,000 participants aged 40–69 years when recruited in 2006–2010.^28^ The study has collected and continues to collect extensive phenotypic and genotypic detail about its participants, including data from questionnaires, physical measures, sample assays, accelerometry, multimodal imaging, genome-wide genotyping and longitudinal follow-up for a wide range of health-related outcomes. The UK Biobank study was approved by the National Health Service National Research Ethics Service (ref. 11/NW/0382), and all participants provided written informed consent to participate in the UK Biobank study. Information about ethics oversight in the UK Biobank can be found at <https://www.ukbiobank.ac.uk/ethics/>.

The UK Biobank central team performed genotype calling and imputation. Genotyping was performed using the Affymetrix UK BiLEVE Axiom array on an initial 50,000 participants; the remaining 450,000 participants were genotyped using the Affymetrix UK Biobank Axiom® array that genotyped ~850,000 variants.^29^ Genotypes were imputed by the UK Biobank central team using IMPUTE2 software, using UK10K haplotype reference panel merged together with the 1000 Genomes Phase 3 reference panel. Imputed variants included in the analysis were those with imputation R2>0.7 and MAF>0.5%.

Based on QC metrics provided by UK Biobank, samples were removed if they were non-European, exhibited gender mismatch, excess relatedness, heterozygosity or missingness > 5%, and individuals that were part of the UKB White British ancestry subset were extracted. We then removed additional individuals with low call rates (<98%) in well-called (>90%) markers, reducing the number of samples to 402,689.

Acne cases were selected based on presence of an acne code in linked primary care diagnosis (see Mitchell et al. 2022), hospital episodes or within the self-report questionnaire (all codes in table below). GWAS was run in the UK Biobank white British subset (including related individuals) using REGENIE, with 5,297 cases and 397,392 controls. using the genotyping array, sex and the first 3/5 ancestry principal components as covariates.

| **ICD9 codes**  **(UKB field 41271)** | **ICD10 codes**  **(UKB field 41270)** | **Self-report codes (UKB field 20002)** | **Selected V2 read codes (UKB field 42040)** | **Selected CTV3 read codes (UKB field 42040)** |
| --- | --- | --- | --- | --- |
| 706.0  Acne varioliformis | L70.0  Acne vulgaris | 1548  acne/acne vulgaris | M2610  Acne vulgaris | M2610  Acne vulgaris |
| 706.1  Other acne | L70.1  Acne conglobata |  | M261.  Other acne | M261.  Other acne |
|  | L70.2  Acne varioliformis |  | M261X  Acne, unspecified |  |
|  | L70.3  Acne tropica |  | M261A  Pustular acne | M261A  Pustular acne |
|  | L70.4  Infantile acne |  | M261z  Other acne NOS | M261z  Other acne NOS |
|  | L70.5  Acne excoriee |  | M2616  Cystic acne | M2616  Cystic acne (vulgaris) |
|  | L70.8  Other acne |  | Myu6F  Acne, unspecified | Myu6F  Acne, unspecified |
|  | L70.9  Acne, unspecified |  | Myu68  Other acne | Myu68  Other acne |
|  |  |  | 2FG5.  Acne scar | X75uJ  Acne scar  Post-acne scarring |
|  |  |  |  | XE1BO  Acne vulgaris |
|  |  |  |  | X50AA  Acne excoriee  Acne excoriee des jeunes filles  Excoriated acne |
|  |  |  |  | X50A4  Acne |
|  |  |  |  | X50A6  Comedonal acne |
|  |  |  |  | X50A7  Nodulocystic acne |
|  |  |  |  | XE1Cy  Acne vulgaris: [comedo]  Acne vulgaris  Pustular acne  Vulgaris - acne |
|  |  |  |  | m7...  Acne - oral preparations |

**10. ESTONIA BIOBANK**

The Estonian Biobank (EstBB) is a population-based biobank with more than 200,000 participants (https://genomics.ut.ee/en/content/estonian-biobank). The EstBB project is being conducted according to the Estonian Gene Research Act, and all participants have signed a broad informed consent form.^30^ Acne cases (*n* = 30,193, current mean age 34.44 (SD = 9.53) years, 74.5% females) were defined by the ICD-10 codes L70.0 (acne vulgaris), L70.1 (acne conglobata), L70.5 (acne excoriata), L70.8 (other acne), L70.9 (acne, unspecified). Data were collected from the biobank’s health database which is regularly linked with national electronic health registries (including hospital databases). In addition, upon recruitment participants completed a questionnaire, including data about their health status, lifestyle and diet - 3.7% of acne cases (1,121 out of 30,193) are assigned only on the basis of mentioned medically assisted questionnaire and the rest 96.3% of acne diagnosis are coming from the questionnaire and/or electronic health records. EstBB participants with no diagnosis of L70, L71, or L73 served as controls (*n* = 94,694, current mean age 40.32 (SD = 9.42) years, 60.0% females). All individuals are of European descent.

The EstBB samples were genotyped at the Core Genotyping Lab of the Institute of Genomics, University of Tartu, using Illumina global screening arrays v1.0, v2.0 and v2.0_EST. Individuals whose sex defined based on X chromosome heterozygosity did not match the sex recorded in phenotype data were excluded from the analysis. Before imputation, variants were filtered by call rate < 95%, Hardy–Weinberg equilibrium *P* < 1 × 10^-4^ (autosomal variants only), and minor allele frequency < 1%. Prephasing was performed using Eagle v2.3 software,^31^ and imputation was performed using Beagle v.28Sep18.793^32^ with the use of an Estonian population–specific imputation reference panel built from 2,297 whole-genome sequencing samples.^33^

**11. QSKIN**

QSKIN is a large volunteer community sample from Queensland, Australia that was invited for study participation through random draw from the electoral role. Participants answered several questions about their experiences with acne. In this study, we defined cases on the question “*Many people suffer from acne during their lives. How much acne did you have when you were a teenager?*” Participants that responded *Never* were coded as controls, while participants that responded *Mild*, *Moderate* or *Severe* were coded as cases. This resulted in an overall sample size of 8650, 3607 cases and 5043 controls. Genotyping was conducted using the Illumina Infinium Global Screening Array platform. Prior to imputation, a common set of high QC markers between the different genotyping batches was obtained. Marker exclusion criteria included: unknown or ambiguous map position and strand alignment in a BLAST search, missingness >5%, p(HWE test)< 10^-6), MAF<1%, GenTrain score <0.6. The Michigan imputation server was used to impute the genotypes using the HRCr1.1 as a reference panel. Individuals were excluded based on high missingness (missing rate > 3%), inconsistent (and unresolvable) sex, or if deemed ancestry outliers from the European population (6 sd deviations from the first two genetic principal components from 1000 Genomes). Imputed genotype dosages were used for the analyses. GWAS was carried out in SAIGE (v0.44) in R 3.6.1 using a generalised linear mixed model to account for population stratification, cryptic relatedness and unobserved genetic confounding. The GWAS was further adjusted for age, sex, genotyping batch and the first ten principal components.

**12. Genes and Health**

Genes & Health is a community based, long-term study of health and disease in British Bangladeshi and British Pakistani people in east London. Participants were recruited at mosques, libraries, GP surgeries and outpatient clinics by bilingual health researchers. Participants all give consent to electronic health record linkage and donate saliva samples for genetic investigations.

50,000 samples from volunteers were genotyped on the Illumina Infinium Global Screening Array v3.0 (with an additional 46 662 Multi-Disease variants) and imputed with the TOPMED-r2 imputation panel. Data were combined across primary care, NHS digital and hospital records to derive the first occurrence of codes mapping onto the three digit ICD10 code for acne. Participants were defined as cases if they had an ICD10 code and controls were those participants without an ICD10 code for acne. After quality control exclusions, this totaled 4,094 cases and 37,935 controls. Association testing was performed with the Regenie software package (version 3.2.2).

**13. eMERGE African American Cohort**

An acne GWAS was performed in a cohort of genetically defined African Americans using data from the electronic MEdical Records & GEnomics (eMERGE-III) consortium.^34^ This NIH-funded initiative has created a large electronic biorepository that contains more than 30 million observations from electronic health records (EHRs) linked to genome-wide genotype data for 102,138 research participants. This research protocol was approved by relevant institutional review boards. Genetic variants have been imputed with the Haplotype Reference Consortium panel.^3^ We first used a subset of 40,156 autosomal single nucleotide polymorphisms (SNPs; minor allele frequency >.05) and in linkage equilibrium to estimate population ancestral components and construct a cohort of research participants with African genetic ancestry.^35^ We next used a diagnosis code billing code to identify acne cases and controls, whereby patients with two or more records of ICD9 code 706.1 were classified as cases and those without any record of 706.1 were classified as controls. In order to control for population stratification, principal components (PCs) were estimated from ancestry informative SNPs.^3^ Finally, association tests were run using PLINK v1.9,^8,36^ adjusting for sex and the first five PCs.

**14. Generation R**

The Generation R study, which began in 2002 in Rotterdam, the Netherlands, is an ongoing, population-based prospective study. Its main goal is to uncover early genetic and environmental factors that influence both typical and atypical growth, development, and health from fetal life through childhood and young adulthood. The study enrolled children and their parents starting from the prenatal period, with a focus on understanding how early genetic and environmental factors impact health and the risk of diseases later in life. At age 13, participants had standardized 3D facial photos taken from both front and side views. Two trained evaluators independently assessed these photos using the Global Evaluation of Acne Severity (GEA) score. Acne severity was categorized into three levels: (almost) clear (GEA 0-1), mild (GEA 2), and moderate-to-severe (GEA 3-5). Acne was considered present at GEA scores of 2 to 5. The final study sample included 4,561 participants, and genotyping was performed using the Illumina 670K platform at the Human Genomics Facility (HuGe-F) of the Genetic Laboratory of the Department of Internal Medicine.

The PGS analysis was done using imputed datasets from Generation R using the 1000Genomes (1000G_PhaseIIIV.5) reference dataset. There were two datasets because the data were genotyped using two different arrays, namely	IlluminaHap 610	and GSA array. Details of the genotyping and QC can be found in Medina-Gomez et al.^37^ Quality control was done on the imputed datasets. (details can be found at <https://github.com/inDEPTHlab/PRS/blob/main/vcf_conversion/readme_bed_rds_files.md>). We performed GWAS PGS regressions per cohort on Acne vulgaris severity with the list of SNPs provided by the lead investigators and using the function score from plink. (<https://www.cog-genomics.org/plink/1.9/score>)

2. **SUPPLEMENTARY FIGURES**

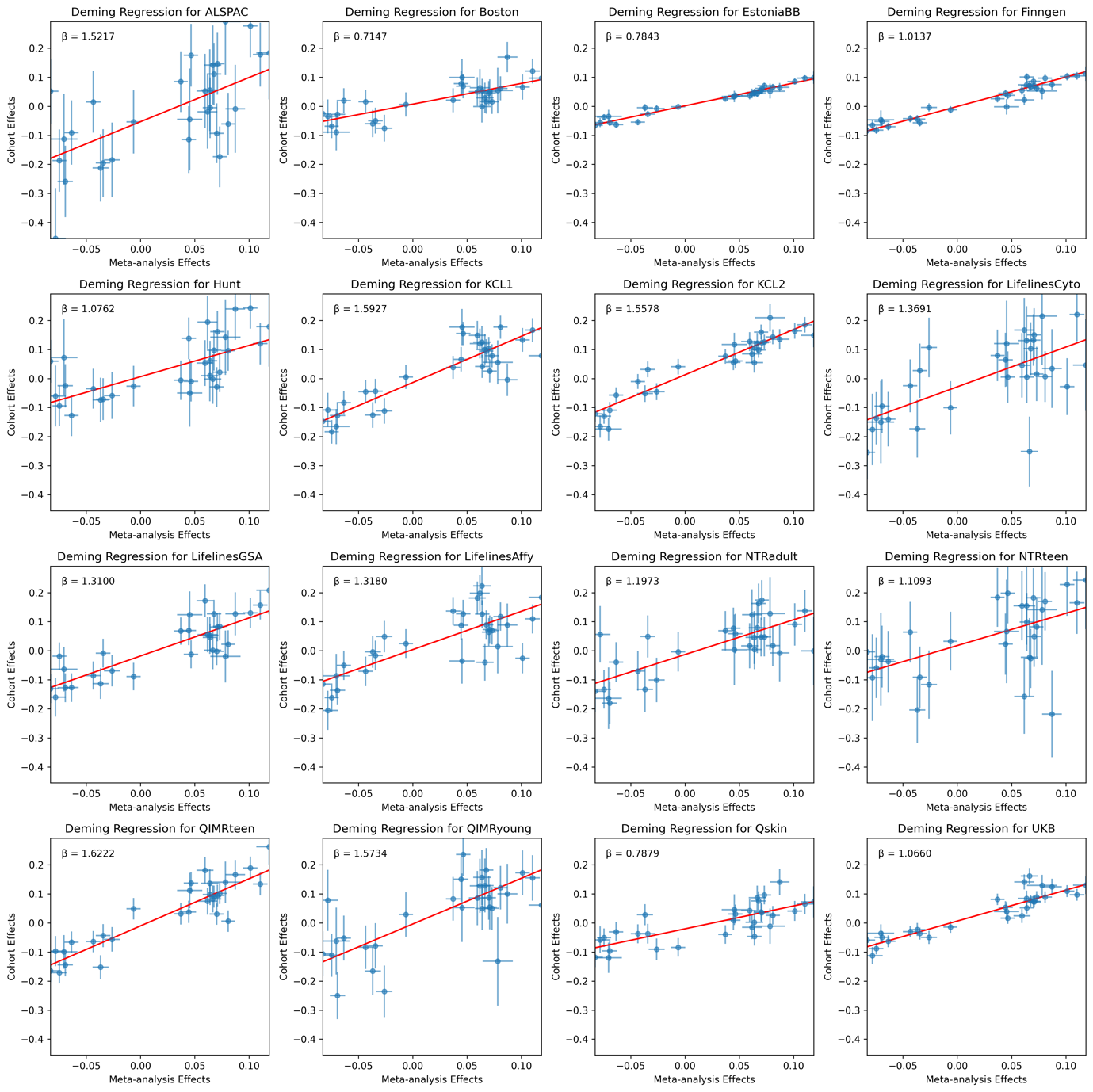

**Supplementary figure 1.** Deming regression analyses for 16 individual GWAS cohorts on the effect size of 30 established acne loci index SNPs compared to a meta-analysis of the same SNPs. Each subplot displays the cohort-specific effects on the y-axis against the meta-analysis effects on the x-axis, with error bars representing standard errors. The red line represents the Deming regression fit, and the slope ($\beta$) value, indicating the degree of concordance between cohort and meta-analysis effects, is annotated within each plot.

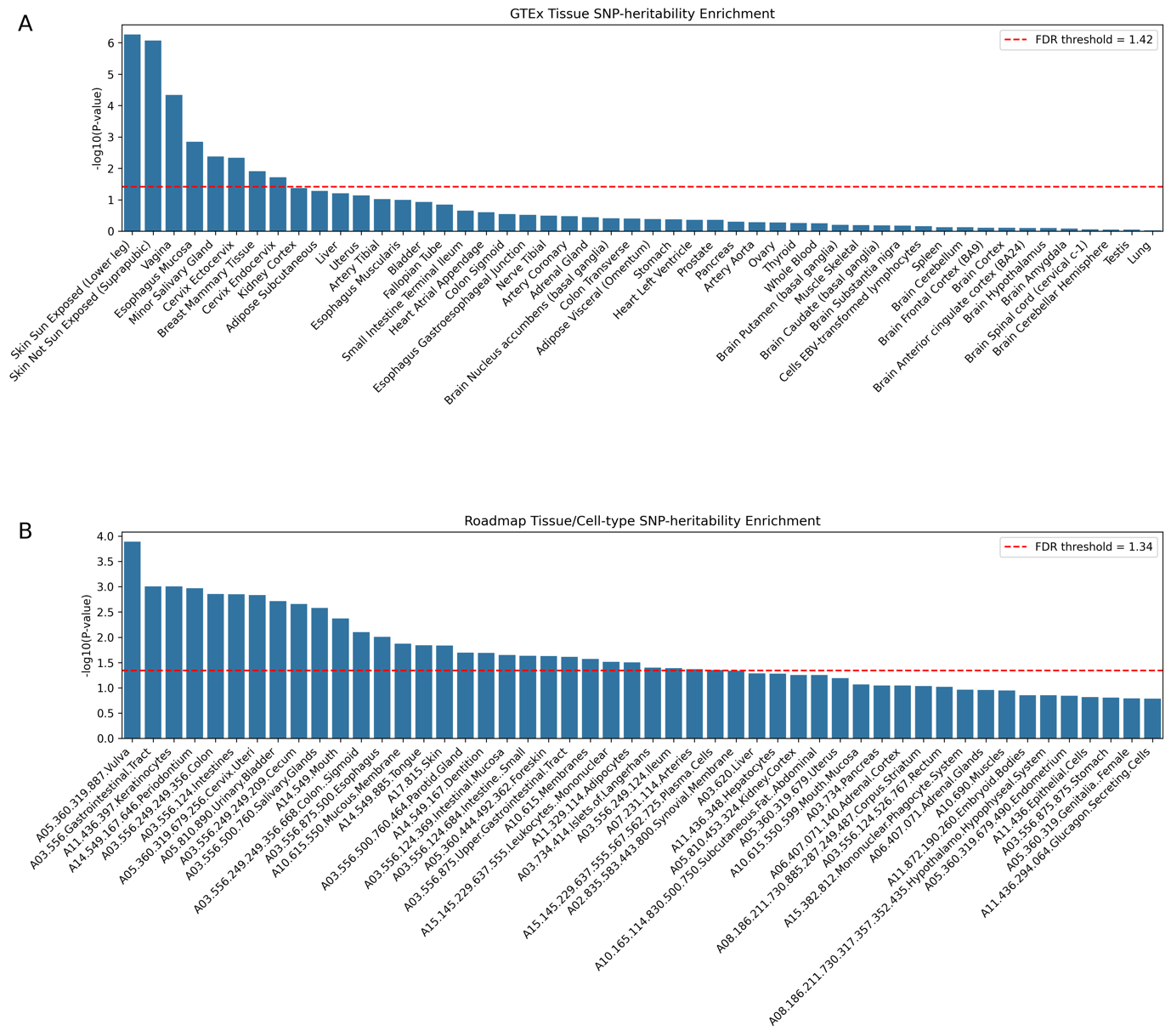

**Supplementary figure 2.** Enrichment analysis for tissues and cell types. Partitioned SNP-heritability for acne by gene expression annotations for GTEx tissues **(A)** and Roadmap tissues and cell-types **(B)**. The x-axis labels denote the specific tissues or cell-types, while the y-axis shows the corresponding -log10(P-value). The plots include the top 50 significantly enriched tissues or cell-types based on their -log_10_(P-value). In each panel, the red dashed line represents the -log10 of the false discovery rate (FDR) threshold, calculated using the Benjamini-Hochberg method.

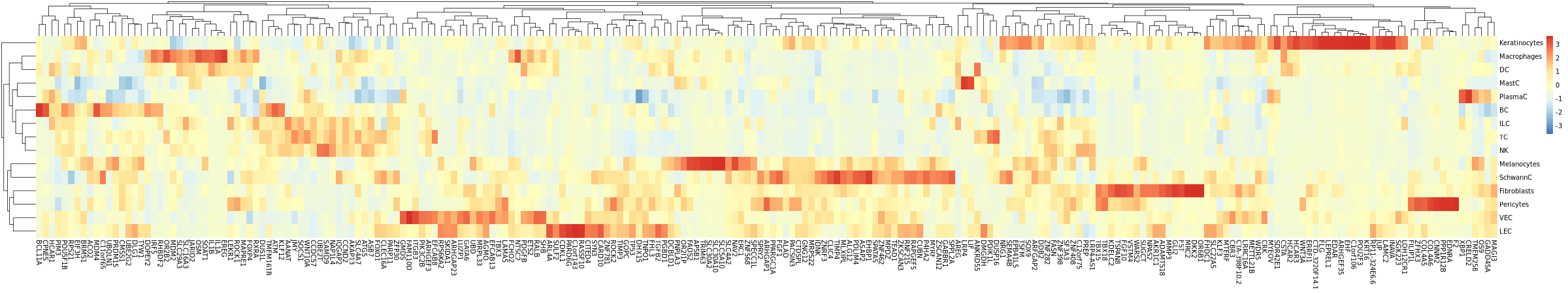

**Supplementary Figure 3.** Analysis of single-cell RNA sequencing (scRNA-seq) data from facial tissue, highlighting the expression levels of prioritised acne genes across different cell types. The heatmap shows scaled gene expression levels, where each row represents a distinct cell type: Keratinocytes, Macrophages, Dendritic Cells (DC), Mast Cells (MastC), Plasma Cells (PlasmaC), B Cells (BC), Innate Lymphoid Cells (ILC), T Cells (TC), Natural Killer cells (NK), Vascular Endothelial Cells (VEC), Lymphatic Endothelial Cells (LEC), Schwann Cells (SchwannC), Pericytes, Fibroblasts, and Melanocytes. Each column corresponds to a acne GWAS gene, with colours indicating the scaled mean expression within each cell type.
